# Epigenetic patterns and methylation-based models for robust outcome prediction in osteosarcoma

**DOI:** 10.1101/2025.11.19.25340108

**Authors:** Joshua J. Bowers, Nikhil Ramavenkat, Christopher E. Lietz, Miya Hugaboom, Aditya Madupur, Edwin Choy, Gregory M. Cote, Hang Lee, Martin J. Aryee, Dimitrios Spentzos

**Author notes:** **Corresponding author:**Dimitrios Spentzos, 55 Fruit Street, Boston, MA 02114.

## Abstract

Osteosarcoma (OSA) is an aggressive bone malignancy primarily affecting children and young adults. Long-term survival has remained unchanged for at least three decades, and no robust molecular prognostic factors have emerged that might help improve this outcome. The complex genomic architecture and genetic heterogeneity of OSA have constrained the identification and validation of such molecular predictors. Recent work has shown that DNA methylation is associated with outcome, yet clinically applicable models building on these findings have not been developed. To address this unmet need, we developed and validated region-based methylation models for individualized risk prognostication and treatment response prediction at the time of diagnosis. These findings were further contextualized through an unsupervised exploration of methylation features, showing an association between widespread global hypermethylation and unfavorable outcomes. The prognostic effect of methylation was independent of known confounding factors. Additionally, an examination of methylation-derived epigenetic age provided orthogonal information with significant association to outcome. Finally, clustering phenotypes were compared between methylation and several other genomic layers (mRNA, miRNA, and copy number variation) and showed little overlap with one another. Together, these findings establish a foundation for unique methylation-based outcome stratification at the time of diagnosis and underscore the need for continued investigation toward clinical application.

## INTRODUCTION

Osteosarcoma (OSA) is the most common primary malignant bone tumor with a predisposition to affect children and young adults and remains a leading cause of cancer mortality in pediatric patients^1,2^. Despite the use of multi-agent MAP chemotherapy (methotrexate, doxorubicin, and cisplatin) coupled with surgical resection^3,4^, long term survival rates have plateaued for at least three decades^5–9^. Outcomes remain especially poor for patients who present upfront with metastatic disease, or those whose tumors are found to be chemoresistant upon primary treatment. A key driver of this prolonged clinical stagnation is the failure of the traditional single-gene mutation biomarker discovery paradigm. While precision oncology has found a great deal of success in the management of other malignancies through the discovery of recurrent and actionable genetic mutations and genomic targets, this classical approach has consistently fallen short in OSA^10–13^.

Early and accurate prognostication is not yet possible as there are no established outcome-based biomarkers at the time of diagnosis. Pathology-based assessment of response to chemotherapy, which is the only widely accepted prognostic factor, can only be obtained following exposure to preoperative chemotherapy, at the time of surgical resection. Further, this histological marker has not resulted in new avenues for treatment that may improve the outcome of patients with poor prognosis, as shown in the negative results of the EURAMOS trial^8,14,15^. This is compounded by the high and often early metastatic potential of the disease. Historically, prior to the implementation of neoadjuvant or adjuvant chemotherapy, when amputation was the only treatment for localized tumors, about 80% of patients still developed later metastases. These factors highlight the need to develop molecular (as opposed to simply histologic or clinical) biomarkers that anticipate long-term outcome and may open new avenues to inform treatment strategies at the time of diagnosis.

The genome of OSA tumors is often dominated by complex structural rearrangements resulting in broad copy-number alterations (deletions, amplifications), numerous other structural variants, and an overall picture that has been called “genomic chaos”^16–21^. It is thought that these rearrangements are brought about by massive mutational events like chromothripsis and chromoplexy^17,22,23^. The genomic landscape can be further complicated by whole-genome duplication, plausibly resulting from a stress response to significant loss of copy number and resulting loss of heterozygosity^24^. Notably, emerging evidence suggests that a large portion of the genomic complexity is established early in tumorigenesis rather than occurring from ongoing chromosomal instability^25^. In parallel, alterations in DNA methylation are detectable early and appear to co-occur with these architectural changes^26,27^.

Within this unique genomic architecture, several characteristic recurrent mutations have been identified including alterations in *TP53*, *RB1*, *MYC*, and *ATRX*^16,17,28–32^. However, apart from *MYC* which has only recently shown a convincing association with outcome, none of these mutations have demonstrated consistent prognostic or predictive value^33–35^.

Given the complex structural heterogeneity and the limited yield of single-gene biomarkers in OSA, recent expert consensus in the field has emphasized higher order biomarker work, with potential emphasis on epigenetic regulation^11,12,36^. Early evidence has shown that DNA methylation patterns are associated with survival and response to chemotherapy^37,38^. Across many other cancers, methylation patterns often display a coupling of global hypomethylation and focal hypermethylation of specific oncogenic loci, a pattern that has been linked with disease progression^39,40^. By contrast, in OSA, the effects of global methylation may be less straightforward as suggested by the emerging strong association between global hypomethylation and positive outcomes^37^. Significant gaps still exist in understanding the role of the DNA methylome in OSA and its effect on tumor outcome.

Leveraging the NCI (National Cancer Institute) Therapeutically Applicable Research to Generate Effective Treatments (TARGET) OSA cohort, we conducted a new methylation-focused study. First, using a region-based methylation approach (variably methylated regions; VMRs), we derive unsupervised subgroups to characterize genome-wide methylation patterns. Region-based approaches exploit the local concordance amongst adjacent CpGs to derive methylation features with greatly reduced dimensionality and improved robustness to CpG-level noise^41,42^. We subsequently build, for the first time, fully supervised and internally validated individualized prognostic and predictive models for long-term survival and response to chemotherapy. Reproducibility and generalizability of both the underlying methylation patterns and supervised models were further externally validated using an independent cohort. Additionally, we investigate epigenetic age, its association with outcome, and whether it provides additional and complementary insights to the global prognostic model. Finally, taking advantage of the multi-omics data available in the TARGET cohort, in a cross-platform analysis, we assessed whether our DNA methylation-derived prognostic phenotypes are redundant or complementary to phenotypes derived by transcriptomic, miRNA, and copy-number patterns. Of note, a recently released international panel consensus framework on OSA biomarkers, identified DNA methylation as a promising biomarker recommending further study for clinical translation, underscoring the promise and timeliness of the results presented in this study^43^.

## RESULTS

### Patient Cohorts and Datasets

We analyzed two independent, clinically annotated OSA DNA methylation datasets and their association with clinical outcome. The first dataset comes from the NCI TARGET initiative, which is a publicly available resource hosted on the NCI Genomic Data Commons^44^ (GDC, https://portal.gdc.cancer.gov/projects/TARGET-OS). This cohort includes 83 patients with comprehensive clinical annotations. Genome-wide array-based methylation profiling was performed on the TARGET samples using the Illumina Infinium HumanMethylation450 BeadChip array (450k array). The second dataset comprises 15 OSA samples from the Albert Einstein College of Medicine (AECM). Comparable clinical variables were available for this cohort and methylation profiling was performed using a HELP-tagging assay. In subsequent sections, the AECM dataset acts as a validation cohort for the findings derived from the TARGET dataset. Detailed information regarding both cohorts and assays have been previously reported^37,38,44^.

### Variably Methylated Region Analysis in Relation to Clinical Outcome in Osteosarcoma

#### Derivation of variably methylated regions

To reduce redundancy, multidimensionality, and noise inherent to single-site methylation analyses across 450,000+ CpG sites, we processed and analyzed the TARGET cohort methylome by identifying 4,427 genomic regions with high inter-sample variability and intra-region concordance among CpG sites, termed variably methylated regions (VMRs, Supplementary Data 1)^42^. VMRs were derived from methylation β-values generated from raw intensity files and processed using standard techniques as described in Methods. This collection of VMRs is henceforth called the “global VMR profile”. This profile was distributed across all chromosomes, with apparent enrichment on chromosome 13 and depletion on chromosome 9 (Figure 1a). Each VMR averaged roughly 366 base pairs and encompassed about five CpG sites. Additional characteristics of the VMR distribution are provided in Supplementary Data 1.

**Figure 1:**
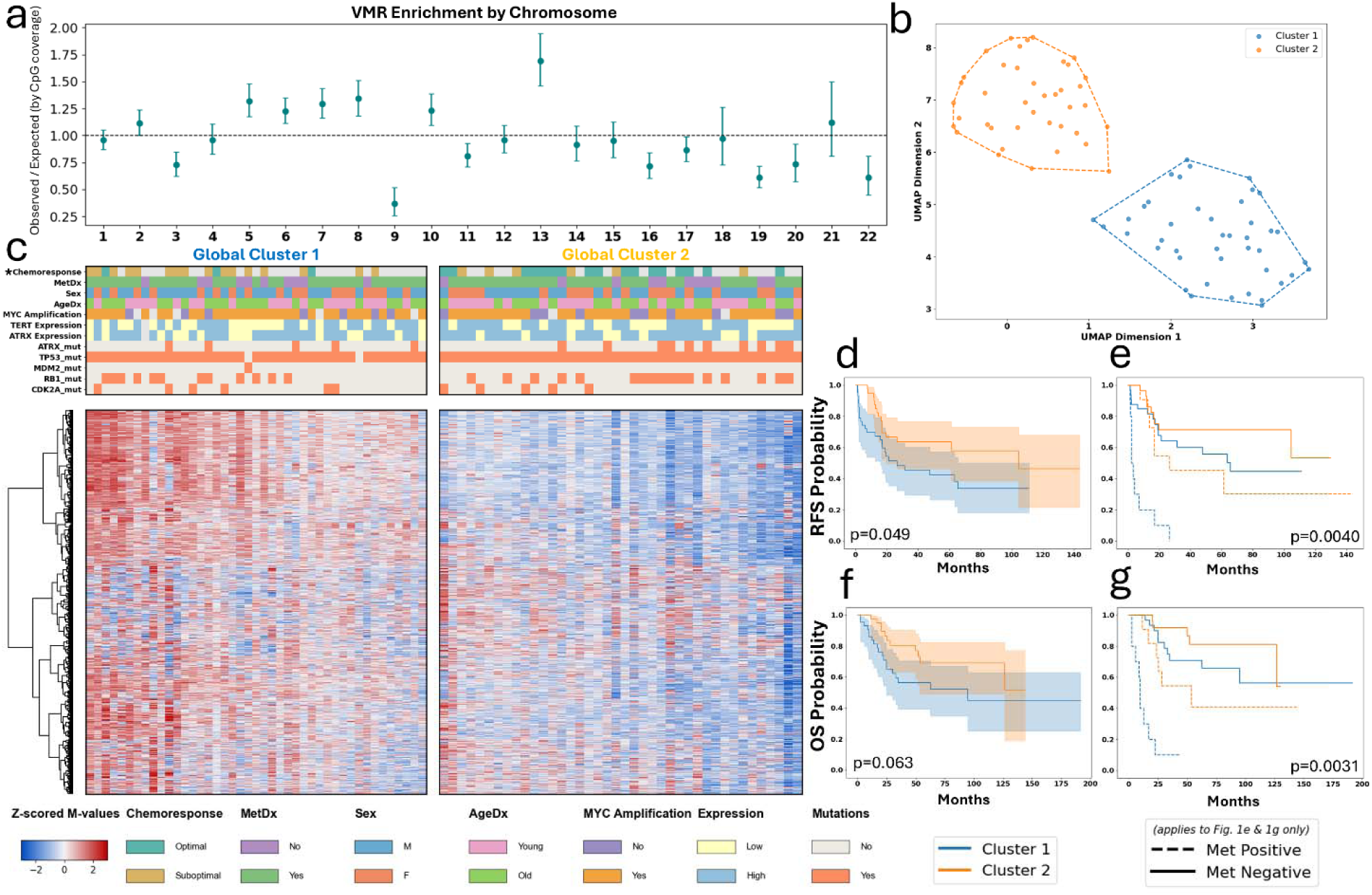
Unsupervised clustering analysis of the TARGET dataset. Figure 1a: Chromosome-level enrichment of global VMR profile, quantified as observed-to-expected counts normalized to Illumina 450k array CpG coverage (95% Poisson confidence intervals). Figure 1b: Unsupervised UMAP projection of TARGET samples by global VMR profile with k-means clustering. Figure 1c: VMR heatmap of global clusters sorted by average methylation. Color bars display clinical and molecular variable distribution. Asterisk denotes the only significant association (χ^2^ p = 6.0 × 10^−4^, OR = 15.11, 95% CI = 3.22-70.84). Figure 1d: RFS analysis of 2-group global VMR UMAP clusters (log-rank p = 0.049). Figure 1e: MetDx stratified RFS analysis of 2-group global VMR UMAP clusters (stratified log-rank p = 0.004). Figure 1f: OS analysis of 2-group global VMR UMAP clusters (log-rank p= 0.063). Figure 1g: MetDx stratified OS analysis of 2-group global VMR UMAP clusters (stratified log-rank p = 0.003).

#### Unsupervised association of the global VMR profile with clinical outcomes

The global VMR profile was analyzed using unsupervised clustering, resulting in two molecularly distinct clusters: cluster 1 (C1, unfavorable outcome) and cluster 2 (C2, favorable outcome) (Figure 1b). Samples in C1 were relatively hypermethylated while samples in C2 tended to be relatively hypomethylated (Figure 1c). A comprehensive summary of all unsupervised survival analyses results is provided in Table 1. Global clustering associated with recurrence-free survival (RFS, log-rank p = 0.049; Figure 1d) and showed a near-significant association with overall survival (OS, log-rank p = 0.063; Figure 1f), demonstrating a link between the hypermethylated subgroup and poor outcome. On an absolute scale, the average beta values across all VMRs differed by 0.15 between C1 (β̅= 0.54) and C2 (β̅= 0.39) (Supplementary Data 2).

**Table 1:**
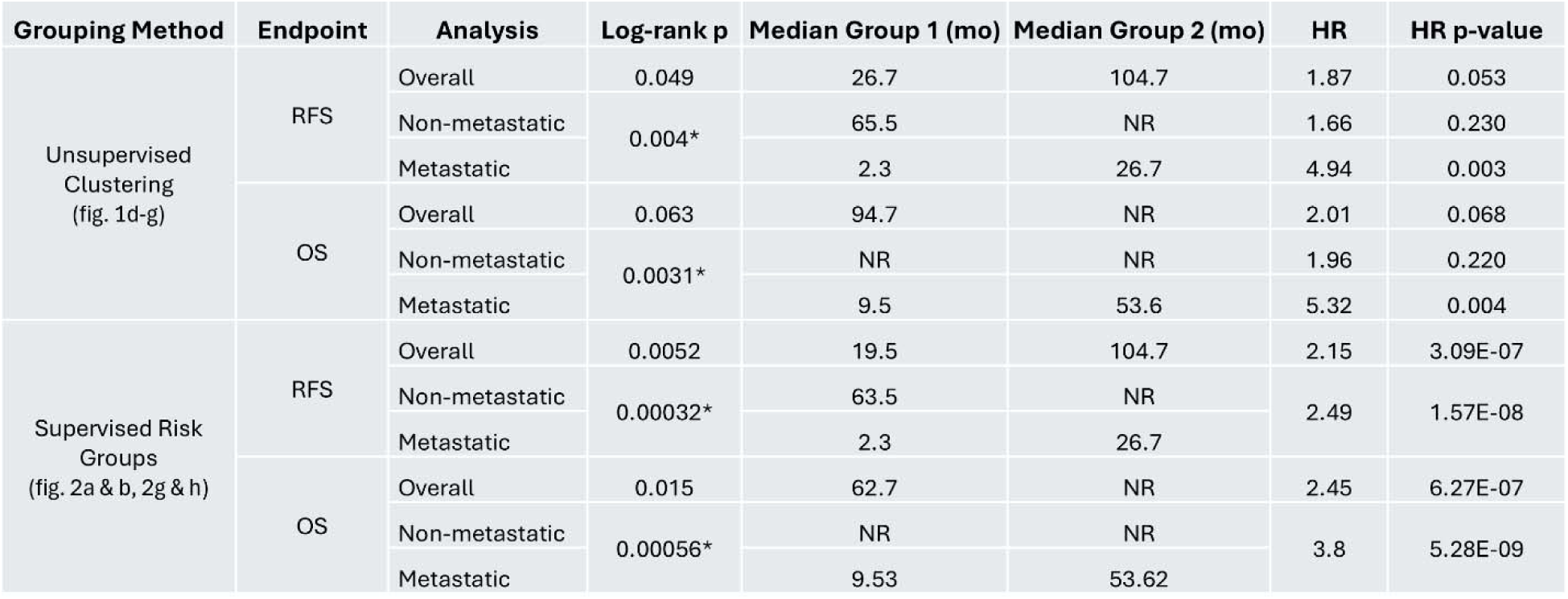
Survival Analysis by Unsupervised Clustering and Supervised Risk Grouping. Survival outcomes for unsupervised clusters and supervised risk groups, with further stratification by MetDx (“*” indicates use of stratified log-rank test). Hazard ratios estimated using binary (unsupervised) and continuous (supervised) Cox proportional hazards models.

After stratifying for metastasis status at diagnosis (MetDx), the association remained and showed stronger statistical significance and effect size for both RFS (stratified log-rank p = 0.004 Figure 1e) and OS (stratified log-rank p = 0.003; Figure 1g). MetDx itself showed no association with average methylation (Supplementary Data 3). In addition, among samples with available annotations, global clusters showed significant association with response to neoadjuvant chemotherapy (chemoresponse; χ^2^ p = 6.0 × 10^−4^, OR = 15.11, 95% CI = 3.22-70.84). Several clinicopathologic variables including sex, age at diagnosis (AgeDx), mutations in several relevant oncogenes or tumor suppressor genes, expression levels of *TERT* and *ATRX*, and copy number alteration of *MYC* were evaluated as potential confounders in relation to global clustering (Figure 1e, Supplementary Data 4); none of which showed a significant association.

Given recent evidence implicating *MYC* amplification as a prognostic factor, we further assessed whether *MYC* copy number alterations confounded the effect of methylation on outcome. We found that the association between global clusters and survival remained largely unchanged across *MYC*-stratified groups. (RFS: stratified log-rank p = 0.059, *MYC* Amplified: HR = 1.91, p = 0.080; Non-Amplified: HR = 1.67, p = 0.533, Supplementary Data 5a; OS: stratified log-rank p = 0.062, *MYC* Amplified: HR = 1.97, p = 0.100; Non-Amplified: HR = 2.95, p = 0.377, Supplementary Data 5b).

### Supervised Analysis of Variably Methylated Regions Reveals Strong Prognostic Risk Stratification

#### Univariable survival analysis of global VMR profile

We next sought to quantify the prognostic signal observed in our unsupervised analysis at the individual VMR level to assess the potential for risk modeling. We first performed Cox Proportional Hazards (CPH) regression analysis of individual VMRs for association with survival. This analysis resulted in 61 VMRs associated with RFS and 1 with OS (q < 0.05, Supplementary Data 6). Due to fewer events and subsequently lower statistical power, the OS stringency threshold was relaxed (FDR q < 0.1), yielding 271 OS-associated VMRs (Supplementary Data 6). When correcting for MetDx, a higher number of significant VMRs was observed (992 and 1464 VMRs, FDR q < 0.05, Supplementary Data 7).

Among the restrictive 61 RFS-associated VMR set, several loci mapped to cytobands containing genes with established relevance to oncogenesis in other tumor types. Several cytobands contained multiple RFS-associated VMRs, including 4q25 (*PITX2*), 7q22 (*GPC2*, *PIK3CG*, *CUX1*), and 14q22.3 (*OTX2*, *OTX2-AS1*), with the most prominent localization at 7q36.3, which contained 13 VMRs mapping to PTPRN2. Notably, several single-VMR cytobands also highlighted canonical cancer-associated genes, including 8q21.3 (*RUNX1T1*), 7q31.32 (*PTPRZ1*), 7q21.2 (*CDK6*), and 10q23.31 (*KLLN*).

To evaluate the biological pathways associated with the RFS associated signature, we performed GSEA on the VMR-mapped gene list. This analysis identified an enrichment or strong trend for enrichment of pathways such as of “Hallmark Estrogen Response Late”, oncogenic RAF signaling, and cellular signatures characteristic of chondrosarcoma stroma and fetal lung neuroendocrine cells. Additionally, we found an overrepresentation of genes associated with specific miRNA regulatory motifs, including targets for MIR5581, MIR6740, and several others. For a full list of all GSEA results, see Supplementary Data 8.

To more specifically elucidate the functional implications of the enriched miRNA targets, we performed a miRNA-centric pathway analysis using miRPathv4.0^45^. These results suggest that the identified miRNAs potentially modulate key oncogenic processes, including ErbB, TGF-β, and MAPK signaling, as well as pathways associated with STK33 activity. A full list of these miRNA-targeted biological pathways can be found in Supplementary Data 9.

#### Supervised risk prognostication

We employed a supervised CPH model to assign risk scores to each sample using a leave-one-out cross validation (LOOCV) approach (Supplementary Data 10). Using all 4,427 VMRs as input at each LOOCV iteration, we recalculated the fold-specific set of univariably significant VMRs and summarized each sample by a signed average methylation score. This score is computed as the mean of those VMR values weighted by the direction of their Cox regression coefficients and serve as input to the supervised CPH model (see details in Methods). Risk scores aggregated under the signed□average cross□validation scheme served as the basis for defining risk groups for subsequent analyses.

Risk scores were dichotomized at the median to define high- and low-risk groups. A comprehensive summary of all supervised survival analyses results is provided in Table 1. Survival analysis demonstrated significance stratification for both RFS (log-rank p = 0.005, Figure 2a) and OS (log-rank p = 0.015, Figure 2g). The absolute difference in average methylation beta values between the two risk groups was 0.15 (high-risk β̅= 0.56, low-risk β̅= 0.41), nearly identical to that observed in the unsupervised analysis. Of note, these risk groups were also concordant with the global unsupervised clusters shown in Figure 1 (Adjusted Rand Index (ARI) = 0.65, χ^2^ p = 9.1 × 10^−13^), supporting the stability of detected molecular phenotypes across unsupervised and supervised approaches alike.

**Figure 2:**
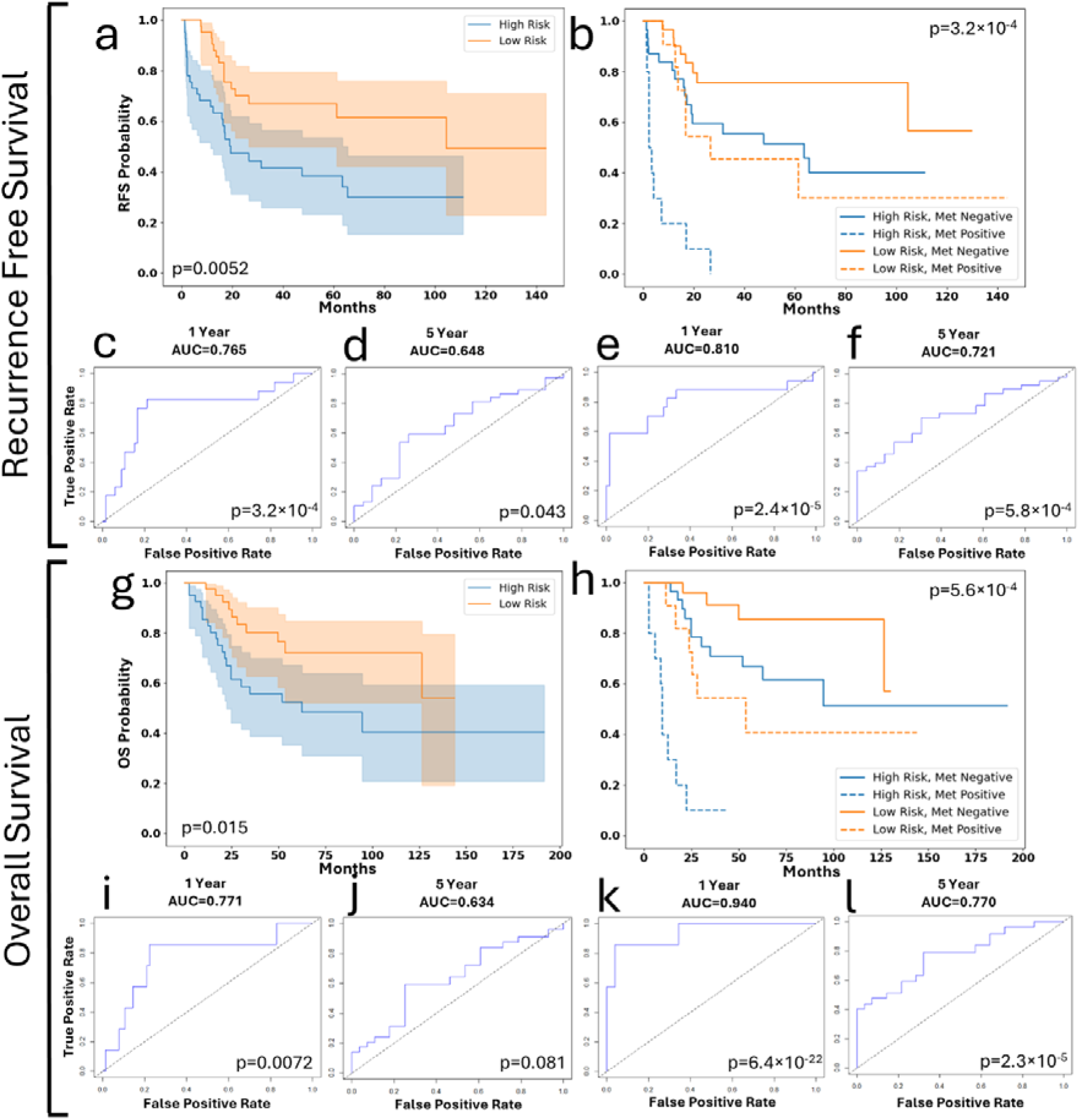
Supervised prognostic survival analysis in the TARGET dataset. Figure 2a: RFS analysis of signed-average methylation risk groups (log-rank p = 0.005). Figure 2b: MetDx stratified RFS analysis of signed-average methylation risk groups (stratified log-rank p = 3.2 × 10^−4^). Figures 2c-d: tdROC analysis of RFS-derived signed-average risk predictions at 1- and 5-year time points (AUC = 0.765, 0.648; p = 3.3 × 10^−4^, 0.043). Figures 2e-f: tdROC analysis of RFS-derived MetDx corrected signed-average risk predictions at 1- and 5-year time points (AUC = 0.810, 0.721; p = 2.4 × 10^−5^, 5.8 × 10^−4^). Figure 2g: OS analysis of signed-average risk groups (log-rank p = 0.015). Figure 2h: MetDx stratified analysis of signed-average risk groups (stratified log-rank p =5.6 × 10^−4^). Figures 2i-j: tdROC analysis of OS-derived signed-average risk predictions at 1- and 5-year time points (AUC = 0.771, 0.634; p = 0.0072, 0.081). Figures 2k-f: tdROC analysis of OS-derived MetDx stratified signed-average risk predictions at 1- and 5-year time points (AUC = 0.940, 0.721; p = 6.4 × 10^−22^, 2.3 × 10^−5^).

Furthermore, risk groups were analyzed against an array of potential confounders, of which only chemoresponse showed an imbalance (Supplementary Data 4). Given the lack of information on chemoresponse for a large subset of patients, a formal multivariate analysis was not feasible. Nonetheless, we also observed that there was still moderate discordance between methylation clusters and chemoresponse (Cramer’s V = 0.67), indicating that risk groups defined by DNA methylation capture prognostic information that is not a simple surrogate of chemoresponse.

In addition, we quantified the association between the signed-average score as a continuous variable and survival. We estimated a Hazard Ratio for RFS of 2.15 (p = 3.1 × 10^−7^, 95% CI = 1.60-2.87). When correcting the association for MetDx, the hazard ratio increased to 2.49 (p = 1.6 × 10^−8^, 95% CI = 1.81-3.42).

Under random label permutation, 97% of iterations yielded a near-zero average number of significant VMRs across cross-validation folds. As such, a stable signed-average predictor could not be constructed for the vast majority of random simulations. This result supports the notion that the observed prognostic signal is improbable under the null hypothesis and likely represents a true biological effect.

The risk groups were further stratified for MetDx, strengthening associations with both RFS (stratified log-rank p = 3.2 × 10^−4^, Figure 2b) and OS (stratified log-rank p = 5.6 × 10^−4^, Figure 2h). Additionally, we applied a time-dependent receiver operating characteristic (tdROC) analysis on the risk scores produced for both RFS (Figures 2c-d) and OS (Figures 2i-j) at 1- and 5-year time points. For RFS, the tdROC area under the curve (AUC) values were 0.765 and 0.648 (p = 3.3 × 10^−4^, 0.043), while the corresponding OS values were 0.771 and 0.634 (p = 0.007, 0.081). Risk scores were then corrected for MetDx (Figures 2e-f, Figures 2k-l), resulting in improved model performance for both endpoints. At 1- and 5-year time points, the MetDx-corrected models achieved AUC values of 0.810 and 0.721 for RFS (p = 2.4 × 10^−5^, 5.8 × 10^−4^), and 0.940 and 0.721 for OS (p = 6.4 × 10^−22^, 2.3 × 10^−5^).

### Supervised Analysis of Variably Methylated Regions Strongly Predicts Response to Neoadjuvant Chemotherapy

#### Univariable chemoresponse analysis of global VMR profile

We next assessed the global profile’s association with chemoresponse in the 43 TARGET samples with available annotations. Univariable analysis identified 952 chemoresponse-associated VMRs (FDR q < 0.05; Supplementary Data 11), only 22 of which overlapped with the 61 VMR survival profile.

#### Supervised chemoresponse predictions

Treating chemoresponse as a binary endpoint, we trained and tested a single-predictor signed-average logistic regression model using a LOOCV approach, emulating the prognostic CPH framework above (Figure 3a, Table 2). The model achieved an AUC = 0.770, (p = 0.001). Given the complexity and broad coverage of the chemoresponse profile (comprising several hundred VMRs), we also tested two alternative methods: L1-penalized logistic regression (Figure 3b, Table 2), and random forest on individual VMR features (Figure 3c, Table 2), both showing largely comparable or stronger performance with AUCs of 0.857, (p = 2.0 × 10^−5^), and 0.744, (p = 0.001), respectively.

**Figure 3:**
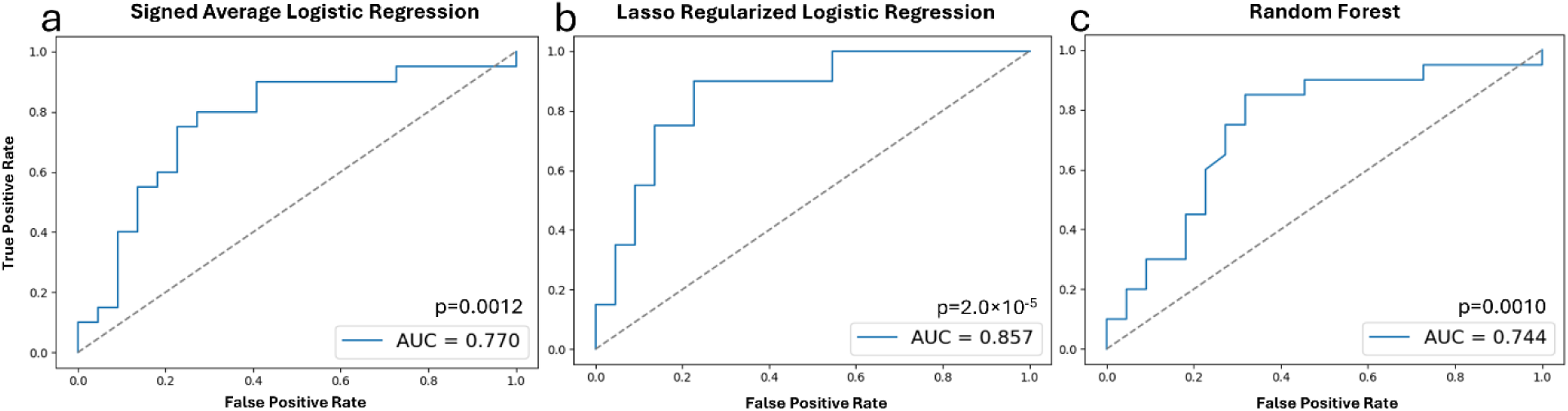
Supervised predictive analysis for Chemoresponse in the TARGET dataset. Figure 3a-c: ROC analysis of (a) signed-average, (b) regularized logistic regression, (c) random forest CR prediction model (AUC = 0.770, 0.857, 0.744; p = 0.001, 2.0 × 10^−5^, 0.001)

**Table 2:** Model Performance Across Clinical Endpoints. Model performances were evaluated using tdROC (survival) and standard ROC (chemoresponse). For RFS and OS, AUC values are shown at 1□ and 5□ year time points using a signed□average risk model, with and without MetDx adjustment. Chemoresponse models include signed-average logistic regression, L1-penalized logistic regression, and random forest methods.

| Endpoint | Model | Time point | AUC | p-value | 95% CI |
| --- | --- | --- | --- | --- | --- |
| RFS<br>(fig. 2c-f) | Signed-average risk score | 1 year | 0.765 | 3.30E-04 | 0.620–0.909 |
|  |  | 5 years | 0.648 | 0.043 | 0.505–0.791 |
|  | MetDx-corrected signed-average risk score | 1 year | 0.810 | 2.40E-05 | 0.666–0.954 |
|  |  | 5 years | 0.721 | 5.80E-04 | 0.595–0.847 |
| OS<br>(fig. 2i-l) | Signed-average risk score | 1 year | 0.771 | 0.007 | 0.573–0.968 |
|  |  | 5 years | 0.634 | 0.081 | 0.483–0.785 |
|  | MetDx-corrected signed-average risk score | 1 year | 0.940 | 6.40E-22 | 0.850–1.00 |
|  |  | 5 years | 0.721 | 2.30E-05 | 0.645–0.894 |
| Chemoresponse<br>(fig. 3a-c) | Signed-average logistic regression | na | 0.770 | 0.001 | 0.616–0.906 |
|  | L1-penalized logistic regression | na | 0.857 | 2.00E-05 | 0.725–0.957 |
|  | Random forest | na | 0.744 | 0.001 | 0.585–0.892 |

Apart from the sparsity intrinsic to L1 regularization, no explicit univariable feature filtering was conducted for the primary chemoresponse analyses. A sensitivity analysis applying feature filtering at two stringencies (nominal p < 0.005 and 0.0025) resulted in negligible differences in model performance. This suggests that chemoresponse methylation markers may reflect a broader cell state as opposed to a narrow mechanism based on a small number of pathways. Comprehensive results for all models and feature filtering stringencies can be found in Supplementary Data 12.

### Prognostic Methylation Profiles and Models are Reproducible in an Independent Validation Dataset (AECM)

#### Mapping and derivation of AECM-VMR set

We tested the robustness and prognostic power of the methylation profiles defined on the TARGET dataset using a publicly available, independent dataset from the AECM. This cohort included 15 OSA samples with methylation characterized by a HELP-tagging assay, a distinctly different methylation assay than the Illumina 450k array. Following the preprocessing steps, we mapped the 4,427 TARGET-derived VMRs on to the HELP-tagging sequence fragments and retained VMRs overlapping with at least one of said fragments. This yielded 3,388 cross-platform VMRs, henceforth called the AECM-VMR set (Supplementary Data 13). Using these VMRs together with the cohort’s corresponding clinical variables, we treat the AECM dataset as an external, independent validation cohort for our findings.

#### Validation of survival analysis

Of the 61 VMRs associated with RFS in the TARGET dataset, 51 mapped to the HELP-Tagging space (Figure 4a). Given the much smaller sample size of the AECM cohort (n=15) and the attrition introduced by cross-platform mapping, univariate associations in the validation set were evaluated at a relaxed threshold of p < 0.10. Under this criterion, 9 and 6 VMRs were univariably associated with 5-year EFS and OS, respectively. Two-group semi-supervised hierarchical clustering using the 51 AECM VMRs yielded a trending association with 5-year OS (χ^2^ p= 0.100, Figure 4a). In a fully supervised manner, we tested a signed-average model, where the sign of the original TARGET CPH model coefficients was applied to the AECM methylation values for these 51 VMRs. Importantly, no information from the AECM dataset was used to train this model. Additionally, AECM did not provide patient censoring information, and thus the prognostic model was tested using logistic regression on a binarized 5-year survival endpoint. The resulting model, trained on the TARGET cohort and directly applied to the AECM cohort, achieved an AUC of 0.860 and 0.906 for 5-year EFS and OS, respectively (p = 0.028, 0.007; Figure 4b-c).

**Figure 4:**
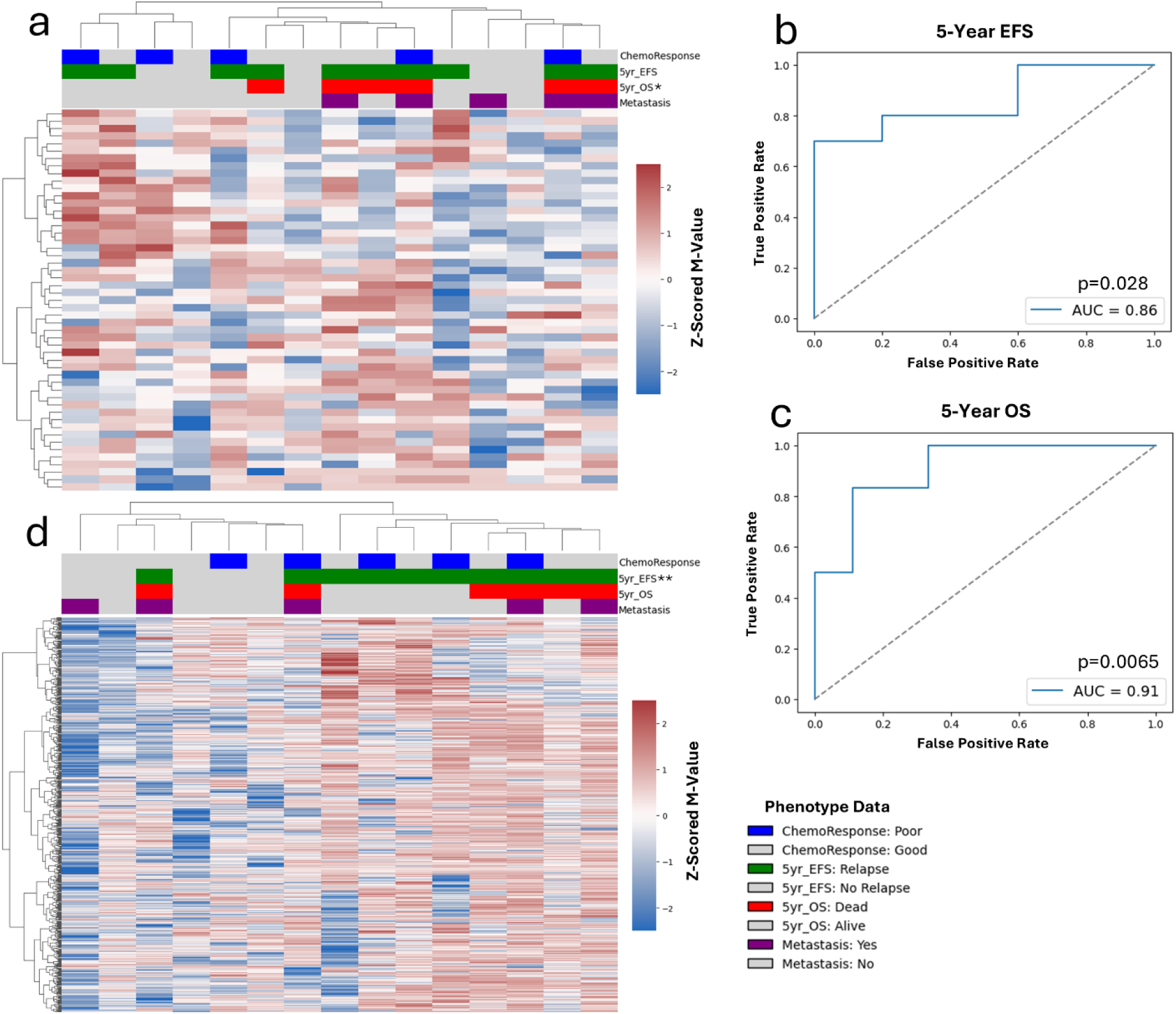
Independent validation analysis in the AECM dataset. Figure 4a: Unsupervised clustering and heatmap of RFS-associated VMRs. Figures 4b-c: ROC analyses of prognostic signed average model trained on TARGET cohort and applied to AECM dataset for 5-year EFS and OS (AUC = 0.860, 0.906; p = 0.028, 0.007, 95% CI = 0.640-1.00, 0.722-1.00). Figure 5d: Unsupervised clustering and heatmap of CR-associated VMRs *Note: * denotes p <= 0.1; ** denotes p <= 0.01.*

Further, of the 952 VMRs associated with chemoresponse in the TARGET dataset, 680 mapped to the HELP-Tagging space (Figure 4d). Semi-supervised hierarchical cluster of those VMRs yielded two groups with a significant difference in 5-year EFS (χ^2^ p = 0.007, Figure 4d). Additionally, methylation patterns strongly differentiate between the two main clusters with hypermethylation being associated with the relatively poor prognosis subgroup, mirroring what was observed in the TARGET dataset.

Given the inherent challenges of cross-platform conversion, the very small number of chemoresponse events (and especially the rarity of chemo-sensitive cases), the reduced sample size of the TARGET training set in chemoresponse analyses, and the large number of univariably-significant chemoresponse VMRs, validation of a fully supervised multi-VMR chemoresponse prediction model in the AECM cohort was not feasible.

### Functional genomic and enhancer-context subset analysis of the global VMR profile

#### Functional subset analysis of global VMR profile

We next investigated whether subgrouping by specific functional genomic regions, called resort types, provided further biologically informed insights into the effects of methylation on survival. The distribution of the global VMR profile by resort-type category can be found in Supplementary Data 14. Unsupervised clustering was applied on each of the resort-type subgroups, where two-cluster solutions were consistently found. Of the four resort-type subgroup clusters, only the open sea region VMRs resulted in a significant RFS association (Supplementary Data 15a-d). However, after stratifying for MetDx, the remaining three resort-type subgroup clusters also became significantly associated with RFS (Supplementary Data 15e-h).

#### Enhancer subset analysis of global VMR profile

Next, we similarly investigated the subset of the global VMR profile that mapped to enhancer elements and examined their association with RFS. To identify tissue- and disease-specific enhancer regions relevant to osteosarcoma, we developed a catalog of enhancers from an independent H3K27ac ChIP-seq dataset^46^. Mapping these enhancers to the global VMR profile, we identified 225 enhancer-linked VMRs. Unsupervised clustering was applied, yielding a two-cluster solution. A trending but non-significant association with RFS was observed (log-rank p = 0.140; Supplementary Data 15i), which became significant after stratifying by MetDx (stratified log-rank p = 0.009; Supplementary Data 15j).

Additionally, the enhancer subgroup was further subdivided by resort-type status, mirroring the analysis done on the global VMR profile. Unsupervised clustering again yielded clear two-cluster solutions across all subgroups. Among enhancer-linked VMRs, those located in CpG island, shores, and open sea regions showed significant associations with RFS (log-rank p = 0.010, 0.029, 0.001, respectively; Supplementary Data 15k-m), while shelf-localized VMRs were observed to be insignificant, yet trending (log-rank p = 0.087; Supplementary Data 15n). After stratifying for MetDx, all regions-specific enhancer subgroups remained or became significant (stratified log-rank p = 0.050, 2.1 × 10^−5^, 0.017, 0.031, respectively; Supplementary Data 15o-r).

### Epigenetic Age Effectively Predicts Both Survival and Response to Chemotherapy

#### Horvath clock, epigenetic age, and association with outcome

Prior work within cancer biology links epigenetic age acceleration with increased cancer risk^47^. This prompted the question of whether assessing epigenetic age via a predefined methylation clock could provide further insights into tumor behavior and aggressiveness. We used the previously published Horvath’s epigenetic clock^48^, a tissue-agnostic algorithm, to assess both survival and chemoresponse in the TARGET dataset and evaluated whether the resulting epigenetic-age stratification was overlapping or orthogonal to that of global methylation clustering (Figure 1).

We examined the Horvath clock output to calculate epigenetic age and subsequently ΔAge (epigenetic – chronological) for each sample (ΔAge distribution shown in Figure 5c). Most samples showed age acceleration, with only 6 of 83 exhibiting deceleration. Additionally, epigenetic and chronological ages were weakly and non-significantly associated (Pearson r = 0.20, p = 0.08).

**Figure 5:**
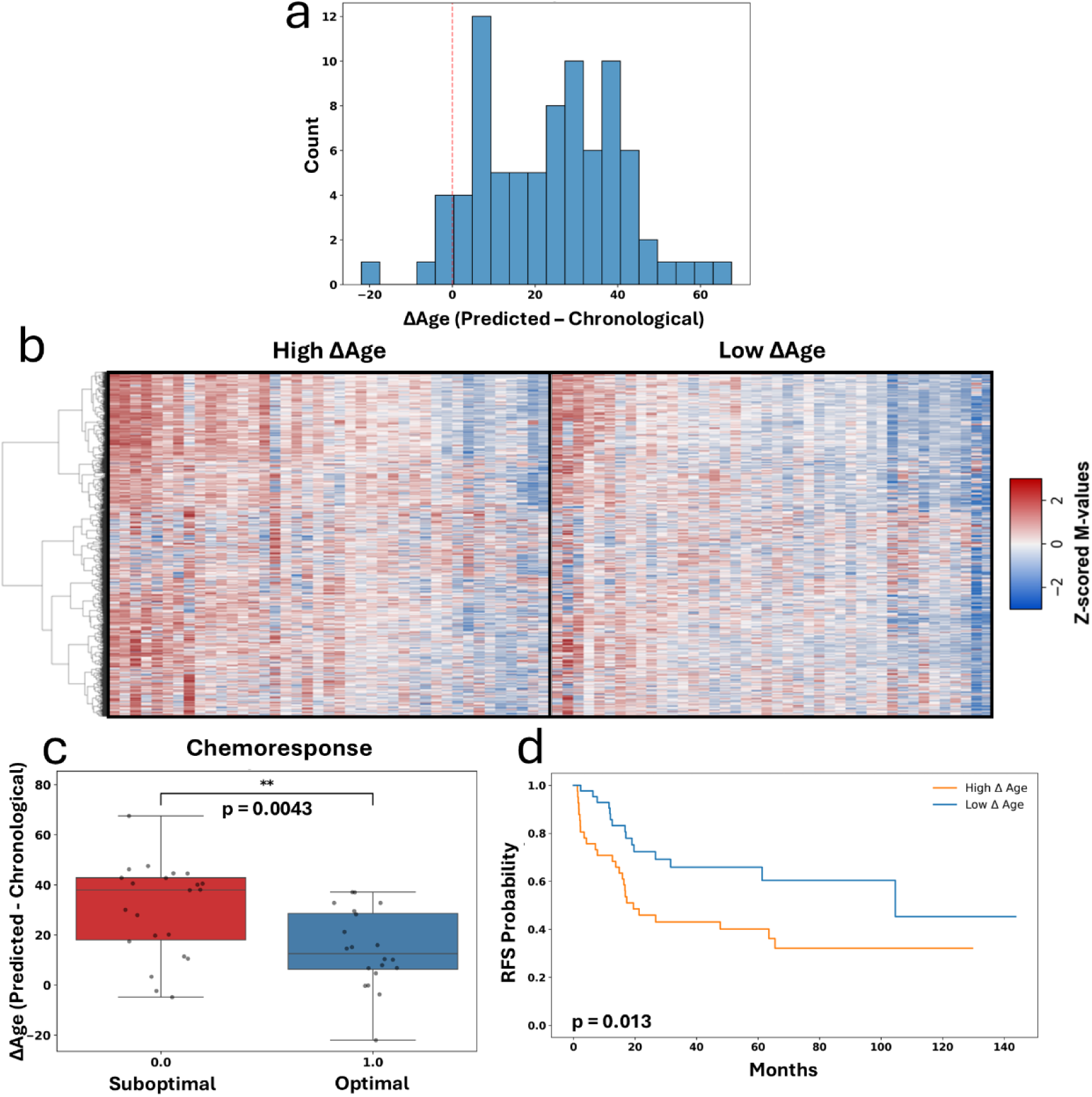
Epigenetic Clock Analysis in the TARGET dataset. Figure 5a: Distribution of ΔAge across cohort as predicted by epigenetic clock (ΔAge = Predicted-Chronological). Figure 5b: VMR heatmap of median-split ΔAge groups sorted by average methylation (Mean: High ΔAge = 0.009, Low ΔAge = −0.582; t-test p = 4.4 × 10^−4^). Figure 5c: Association analysis between ΔAge and chemoresponse (t-test p = 0.004, median ΔAge difference between Suboptimal and Optimal treatment response = 25.5 mo.). Figure 5d: RFS analysis of the cohort split at median ΔAge (log-rank p = 0.013).

ΔAge was positively associated with chemoresponse (t-test p = 0.004; median ΔAge difference = 25.49 months; Figure 5d). When dichotomized at the median, ΔAge was also significantly associated with RFS (log-rank p = 0.012, Figure 5e). Consistent with this, CPH analysis showed that increasing ΔAge was associated with greater risk of recurrence (HR = 1.31, p = 0.005, 95% CI = 1.09-1.59).

#### Complementary information from Horvath clock vs global VMR analysis

We then questioned whether the effect of ΔAge on survival could be associated with or confounding the main effect of the global VMR profile. To this end, we first observed that there was no overlap between the Horvath clock and global VMR profile at the CpG level. We then examined the agreement between the median-split ΔAge groups and our global methylation clusters (Figure 1). Although the two classifications were significantly associated (χ^2^ p = 2.8 × 10^−4^), agreement beyond chance was weak (ARI = 0.168), suggesting that they share only limited overlap and capture largely distinct biological signal. Finally, we introduced ΔAge as a covariate in the original supervised CPH model and observed no effect on the prognostic value of the global VMR model, with the hazard ratio remaining unchanged and significant (HR = 2.15 to 2.12, p = 3.1 × 10^−7^ to 2.9 × 10^−5^).

### Multi-Omics Comparative Analysis Reveals Non-Redundant Prognostic Information in Relation to Methylation Effect

#### Single-platform prognostic association analysis

In addition to the methylation data, several other omics datasets were available for the TARGET cohort, including mRNA sequencing (RNA-Seq), miRNA qRT-PCR (miRNA profiling), and copy number (CN) information. We evaluated whether these additional genomic and transcriptomic features offer outcome information that may be redundant with, or complementary to, that of DNA methylation.

We first analyzed each platform separately for prognostic discrimination by performing unsupervised clustering on the TARGET cohort (Figures 6a-c). The RNA-seq partition (k = 2, feature filtering = 90%) was significantly associated with RFS (log-rank p = 0.033, Figure 6d), however, no strong association was noted with chemoresponse. Likewise, miRNA profiling (k = 2, feature filtering = 50%) resulted in a near significant association with RFS (log-rank p = 0.069, Figure 6f), and no association with chemoresponse. Segment-level CN data, analyzed using GISTIC2.0, resulted in 68 regions of significant amplification or deletion across samples. These regions, while not associated with RFS and chemoresponse under two-cluster solutions (Supplementary Data 16a), became strongly significant for both when considering higher-order clustering (RFS: three-cluster log-rank p = 0.010 (Figure 6h), four-cluster log-rank p = 0.017 (Supplementary Data 16b); Chemoresponse: three-cluster χ^2^ p = 0.019, four-cluster χ^2^ p = 0.042).

**Figure 6:**
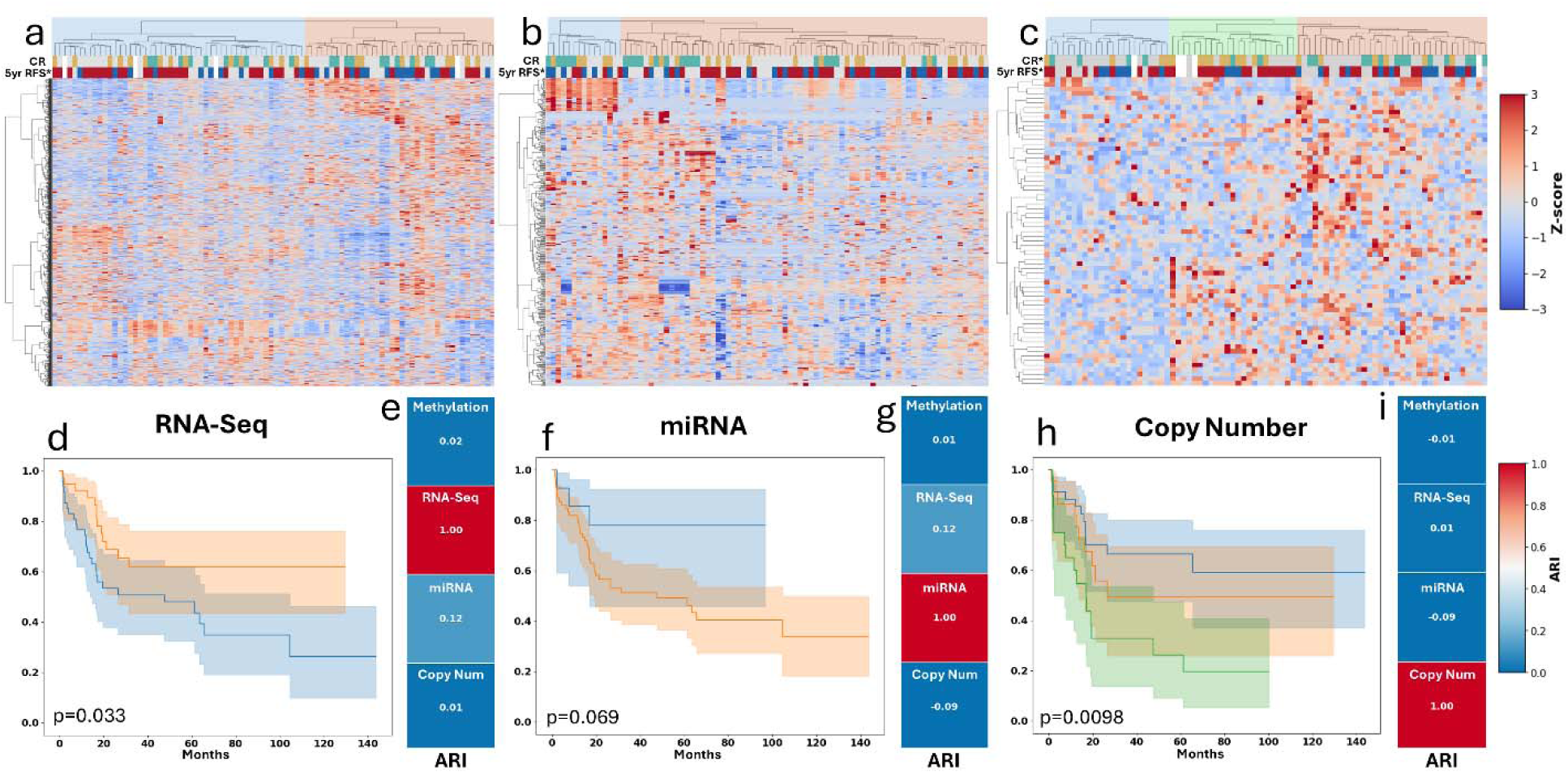
Cross-omics comparison analysis of the TARGET dataset. Figure 6a-c: Unsupervised hierarchical clustering and heatmaps of RNA-seq (a), miRNA-RT-PCR (b), and segment-level copy number (c) platforms. Color bars display chemoresponse (green/brown: top) and 5-year RFS (blue/red: bottom) distribution for each sample. Asterisk denotes association p <= 0.05. Figure 6d-e: RFS (d) and concordance anlaysis (e) for two-group RNA-seq clustering (log-rank p = 0.037; RNA-Seq ARI concordance: Methylation = 0.02, miRNA = 0.12, CN = 0.01). Figure 6f-g: RFS (f) and concordance (g) analysis for two-group miRNA clustering (log rank p = 0.069; miRNA ARI concordance: Methylation = 0.01, RNA-seq = 0.12, CN = −0.09). Figure 6h-i: RFS (h) and concordance (i) analysis for three-group RNA-seq clustering (multi-group log-rank p = 0.001; CN ARI concordance: Methylation = −0.01, RNA-seq = 0.01, miRNA = −0.09).

#### Cross-platform concordance analysis for prognosis

Finally, we assessed concordance across omics platforms to evaluate the extent of overlapping or complementary information (Figures 6e, g, and i). Methylation was clearly distinct and exhibited no agreement with the other three platforms (RNA-Seq ARI = 0.02, miRNA profiling ARI = 0.01, CN ARI = −0.01). Similarly, CN showed minimal concordance with both RNA-Seq and miRNA profiling (ARI = 0.01, ARI = −0.09). And finally, RNA-Seq and miRNA profiling demonstrated slightly more, though still limited, concordance (ARI = 0.12), indicating partial but not complete redundancy.

## DISCUSSION

In the 30 years since the introduction of combined-modality treatment with chemotherapy and surgery in OSA, patient outcomes have plateaued, with 30-40% experiencing relapse after primary tumor treatment. Management of OSA would greatly benefit from the development of molecular biomarkers predicting long-term outcomes at the point of diagnosis. Thus far, more conventional methods of biomarker discovery have been unsuccessful in finding recurrent and prognostic mutations and actionable genomic targets. In fact, the only widely accepted prognostic factor, pathologically assessed tumor necrosis, requires months of chemotherapy before assessment, potentially missing an early window for improved therapeutic development. Additionally, chemoresponse itself has not led to improved treatment outcomes when used in the large international randomized EURAMOS trial^8,14,15^. The complex structural makeup and profound genomic heterogeneity in OSA have obfuscated a clear path toward translational progress in biomarker development. Furthermore, there is now expert consensus in the field, that single gene and single mutation-based outcome biomarkers are unlikely to be useful in OSA^11,12,36^, with c Myc being the only recently recognized exception to this consensus^33–35^.

Although recent work by our group has tied DNA methylation patterns to outcome in OSA^37^, these findings have yet to be translated into robust prognostic and predictive models. In this study, we develop models to predict long-term survival and treatment response using region-based methylation profiles (VMRs). The high dimensionality of methylation data presents a major analytical challenge, as profiling hundreds of thousands to millions of CpGs necessitates aggressive false discovery rate correction. VMRs help overcome this by summarizing redundant methylation across neighboring CpGs into fewer, more stable features that reduce susceptibility to single-CpG noise and improve overall model robustness and generalizability. These VMR-based models are validated both internally and also independently, when possible, based on external data availability. We also perform complementary analyses including an epigenetic age acceleration estimate, providing unique insights into the biology of aggressive OSA. Finally, we perform a multi-platform assessment across RNA-seq, miRNA expression, and copy number variation data, and show that DNA methylation provides largely orthogonal prognostic information to the other types of genomic data. Together, our results establish methylation-based models as practical and promising clinical tools for risk stratification at the time of diagnosis and provide insights into the potential role of DNA methylation in modulating outcome in OSA. We believe that our study is of particular interest and timely, given the newly released consensus on the status and future directions of OSA biomarkers by a leading international osteosarcoma expert panel. In this consensus framework, DNA methylation has been recognized as one of the most promising biomarkers and further work is encouraged for clinical translation^43^.

We have developed methylation-based models for individualized patient risk prognostication and treatment response prediction at the time of diagnosis, representing the first time such predictive modeling has been successfully applied in this disease. Both models have been internally validated using cross-validation techniques, resulting in robust performance quantified by ROC and tdROC analyses. These metrics demonstrate strong and significant discriminatory ability across all outcome variables (RFS, OS, and chemoresponse). Permutation testing further supported the stability of the model, as randomization of survival labels failed to consistently reproduce the observed signal. Additionally, we performed independent validation of survival-based models showcasing their external generalizability. Notably, the two cohorts used in this analysis provided methylation data generated from distinct assay technologies, namely a methylation microarray versus a HELP tagging assay. These robust model performances, despite technical differences and substantial cross-platform attrition, further underscore the reproducibility of our findings.

Importantly, we also have shown that the prognostic power of the supervised model for survival is independent of confounding by known clinical or pathologic prognostic factors, such as metastasis at the time of diagnosis as well as *MYC* amplification, an emerging recently demonstrated prognostic factor. The distribution of these factors was either balanced between the methylation-based risk groups, and/or their inclusion in multivariate models did not affect the prognostic performance of the methylation variable. The only factor that segregated with the risk groups was chemoresponse assessment by pathologic necrosis. Even so, the methylation model does offer non-redundant information compared to chemoresponse. Furthermore, chemoresponse assessment is only available several months after chemotherapy initiation. As such, chemoresponse cannot act as a baseline prognostic tool as it potentially misses a critical window for therapeutic optimization. Moreover, its assessment remains subject to the inherent interobserver variability, and in a subset of cases, for various clinical reasons, chemoresponse is never assessed as patients undergo primary surgery without neoadjuvant chemotherapy.

While a fully independent statistical validation was not possible for the chemoresponse model due to dataset limitations, an unsupervised look into the validation cohort’s chemoresponse-associated VMR profile shows a picture reminiscent of the TARGET chemoresponse profile with global hypermethylation associated with suboptimal chemoresponse. Thus far, genome-wide DNA methylation studies in OSA have been largely associative and lacking clearly defined feature sets suitable for downstream validation. Our models set the foundation for outcome-based prognostication and prediction in OSA, filling a significant translational gap in the field.

Unsupervised analysis of TARGET VMR profile provides potential insights into the biological context of our models. Genome-wide VMR analysis reveals global clusters significantly associated with RFS, OS and chemoresponse. Consistent with our prior CpG-level findings in the TARGET cohort, global VMR clusters show an obvious dichotomy between global hyper- and hypomethylation, with the hypermethylated subgroup being associated with unfavorable outcomes.

Notably, this contrasts the paradigm observed in many other cancers, where global hypomethylation is linked to unfavorable outcomes owing to the improper maintenance of methylation state over successive replications^39,49^. While global hypermethylation is not the dominant pattern associated with poor outcome, it has been linked to unfavorable survival and drug resistance in a few other cancers. In some of these instances, demethylating agents have been effective in sensitizing resistant cells, highlighting a potential significant clinical implication of this epigenetic state^50,51^. Furthermore, *in vitro* treatment of OSA cells with demethylating agents has been shown to inhibit growth by reactivating epigenetically silenced tumor suppressor genes. However, the potential interplay between these drugs and global hypermethylation in OSA remains to be elucidated^26,52–54^. Importantly, several such agents (e.g., decitabine and azacytidine) are already FDA-approved for other hematologic tumors, specifically myelodysplastic syndrome and some leukemias, and therefore represent promising candidates for accelerated therapeutic repurposing in OSA.

Furthermore, univariable associations between methylation features and outcome provide additional insights into potential therapeutic relevance. The chemoresponse-associated VMR profile consisted of a substantial number of largely concordant features. The diffuse nature of this epigenetic signature suggests that chemoresponse may be modulated not by discrete loci, but rather by coordinated changes in cell identity. This could potentially result from the downstream effects of chromothripsis and signify a distinct, global “cell state.” Considering prior observations that cancers with similar global epigenetic motifs can be sensitized by demethylating agents, it is plausible that such agents could synergize with chemotherapy by restoring chemosensitivity in these hypermethylated states. In contrast, supervised survival analysis produced a much smaller subset of associated VMRs, potentially highlighting specific loci that influence long-term outcomes and may therefore require addition of more targeted therapies.

The presence of overt metastasis at the time of diagnosis (MetDx) is a well-established predictor of patient outcome in all solid tumors, including OSA, and serves a crucial role in patient assessment at the point of diagnosis. The evaluation of MetDx showed no significant association with the global VMR clusters, suggesting that they capture some independent biological signal. Indeed, this biological independence was further corroborated by stratified and multivariate analyses, demonstrating that methylation profiling retained prognostic relevance after accounting for MetDx. Interestingly, MetDx-corrected and stratified analyses show evidence of substantially larger effect sizes in patients with metastatic disease. While these findings do not constitute a formal test of interaction, they suggest that effect modification by MetDx could be further explored in future studies.

Global profile VMRs were distributed across the genome in a non-uniform manner, with considerable VMR enrichment and depletion on chromosomes 13 and 9, respectively. Chromosome 13 harbors major tumor-suppressor genes (e.g., *RB1*, *BRCA2*), some of which are implicated in OSA oncogenesis. Additionally, copy number analysis of the TARGET dataset revealed two focal deletion peaks on chromosome 9 (9p24.3 and 9p21.3), potentially accounting for a proportion of the methylation variability. This non-uniform VMR landscape showcases the unique nature of the OSA methylome and suggests that methylation effects align with structural alterations and common functional loci, rather than arising from indiscriminate genomic variation. Furthermore, while proximity of some of the Survival profile VMRs to genes with known role in oncogenesis does not by itself prove causality, they highlight plausible mechanistic candidates that underly survival heterogeneity in OSA.

Resort-type differentiation of global and enhancer-linked VMRs offers insight into the spatial localization and potential functional context of the prognostic signal. Resort-type survival analysis showed a stronger signal in open sea regions to that of the other genomic contexts. At the enhancer level, all regions except shelves showed significant discrimination. Enhancer-linked VMRs localized within CpG islands showed increased discriminatory power than their enhancer-agnostic CpG island VMR counterparts. This plausibly reflects an increase in regulatory potential of enhancers in close proximity to gene promoters. Overall, these findings highlight two key insights into the localization of prognostic signals. First, open sea regions, which are often overlooked in functional annotations, consistently provide strong survival associations in both global and enhancer VMR subsets. Second, enhancer status may provide additional regulatory context, especially in gene dense regions.

We also present the first ever epigenetic age analysis in OSA, as an orthogonal approach to investigating the methylation landscape. Epigenetic clocks estimate biological age based on focal methylation patterns at predefined CpG sites and the resulting epigenetic age values have been linked with increased cancer risk^47^. The TARGET dataset samples overwhelmingly trended toward age acceleration, plausibly resulting from the significant dysregulation of the methylome coinciding with oncogenesis. This assertion is further supported by the lack of strong association between biological and chronological age. Survival analysis linked ΔAge (biological – chronological) to RFS with a significant hazard ratio, indicating that epigenetic age acceleration may contribute to tumor aggressiveness. Groups with high versus low ΔAge revealed significant differences in both RFS and chemoresponse. Epigenetic clock CpGs did not overlap with the global VMR profiles. Furthermore, when comparing these groups with the phenotypes detected by global methylation (Figure 1), a modest, yet functionally minimal, agreement was noted, suggesting that the two approaches capture largely distinct biological signals.

Our comparisons across multiple omics platforms revealed that methylation-based clustering captures a distinct biological signal that is non-redundant with other molecular layers. Although other omics platforms (RNA-seq, miRNA profiling, and CN variation) demonstrated an association with survival, ARI analysis revealed limited concordance among them. Most importantly, methylation phenotypes were distinct from all others indicating that global methylation profiling provides a largely unique signal, independent of transcriptomic, miRNA, and copy number profiles. This independence highlights the potential for complementary modeling approaches that integrate multiple molecular layers to enhance prognostic accuracy and biological insight. Indeed, recent studies have highlighted the potential of integrative multi-omics methods in this setting^33,55^. Nonetheless, the question of whether and to what extent these varying molecular platforms offer overlapping versus orthogonal information, and how they can be best combined synergistically, has yet to be formally addressed. Our current study provides a first, though by no means definitive, analysis of this question.

Our study should be placed in the context of an evolving new focus on epigenetic research in osteosarcoma, which is also becoming a consensus priority in the field^11,12^. Notably, recent work on chromatin accessibility defined prognostically relevant subtypes with translational and therapeutic implications^56^. Additionally, prior work by our group and others produced further insights into a different epigenetic layer, namely miRNA biomarkers and subtypes^57–60^. These observations provide complementary insights with our current study and together set the stage for defining epigenetic patterns as strong biomarkers with mechanistic insights in OSA.

Some constraints of our study should be noted. Given the rare nature of OSA, sample size is a perpetual issue. While this was partially solved with the development of the NCI TARGET multi-omics OSA cohort, external validation still requires an additional sizeable dataset. To this end, our independent validation dataset was small (n=15), which somewhat limits the strength of the conclusions that can be drawn from external validation. Beyond sample size, the TARGET dataset completely lacks annotations for histological subtypes and provides chemotherapy response information for only half of the samples. Thus, formal model adjustment for clinical covariates was limited to metastasis at diagnosis, which nonetheless is the strongest established prognostic factor. Future studies should fully incorporate additional clinical variables, including those featured in published prognostic nomograms^61,62^.

Although VMR-based analyses offer important advantages, VMRs naturally summarize methylation signal across contiguous CpGs, which can potentially overgeneralize and mask important site-level effects. Moreover, the requirement for VMRs to cover at least two CpG sites can result in the exclusion of important single-site CpGs and introduce selection bias towards gene dense regions.

The two cohorts that formed the basis of our study were generated using two different assay technologies: the Illumina 450k microarray and a HELP-tagging assays. Future studies should move toward development of a standardized assay suitable for eventual clinical application. This could conceivably be a custom array with a fixed CpG content. Additional newly available options such as enzymatic methyl sequencing (EM sequencing) or Illumina 5-base sequencing, offer important advantages compared to both conventional bisulfite sequencing and microarrays, and warrant further evaluation.

Future studies should also prioritize the development of a large, independent validation cohort. This would enable rigorous calibration of existing and future predictive and prognostic models, facilitate the definition of clinically meaningful risk thresholds, and provide a foundation for prospective testing in clinical trials. Further, building on our findings that RNA-seq, miRNA, and copy number profiling yield largely non-overlapping yet prognostic signals, integrative modeling approaches that combine different molecular layers could enhance both clustering resolution and model predictive accuracy, reflecting a more comprehensive view of osteosarcoma biology. Additionally, further investigation is needed into the relationship between epigenetic age acceleration and OSA tumor behavior. Finally, future experimental studies should focus on the testing demethylating agents in OSA, especially in the context of enhancing chemoresponse and drug sensitization.

## METHODS

### Study Cohorts and Data Acquisition

Genome-wide DNA methylation data from 83 osteosarcoma (OSA) patients was obtained from the Therapeutically Applicable Research to Generate Effective Treatments (TARGET) initiative^63^. This initiative’s publicly available dataset was hosted via the NCI Genomic Data Commons^44^ (GDC, RRID:SCR_014514, https://portal.gdc.cancer.gov/projects/TARGET-OS). Paired RNA sequencing (RNA-seq), miRNA qRT-PCR (miRNA profiling), and segment-level copy number (CN) data were available for 75 of the 83 methylation samples. The TARGET OSA cohort served as the discovery dataset for all primary analyses.

An independent validation dataset was obtained from the Albert Einstein College of Medicine (AECM)^38^. The AECM cohort consisted of 15 OSA patients with attached methylation information as generated by a HELP-Tagging assay^64^. This publicly available dataset was downloaded from the Gene Expression Omnibus (RRID:SCR_005012, GSE59200). Comparable clinical annotations were available for this cohort, enabling the replication of some of the primary analyses derived from the TARGET discovery dataset.

Additional information regarding both datasets has been previously reported^37^. All data analyzed in this study was publicly available through respective sources. Ethics approvals and informed consent procedures for data acquisition and generation have been described previously^37^. All analyses in the present study were performed in accordance with relevant guidelines and regulations.

### Methylation Data Preprocessing

Methylation profiling of the TARGET cohort was conducted using Illumina Infinium HumanMethylation450 BeadChip (450k array). Raw IDAT files were preprocessed using the minfi package in R (v1.52.1)^65^. Standard preprocessing included normal–exponential out-of-band (NOOB) correction, which uses control probes for background and dye-bias adjustment, was performed^66^. This was followed by functional normalization, leveraging the principal components of control probes to perform between-array correction and reduce unwanted technical variation^67^. Quality control measures were performed, including the removal of probes containing common SNP, probes targeting sex chromosomes, probes targeting cross-reactional sites^68^, and probes where expression does not differ significantly from background noise (>10% of samples with failed probes). CpG annotations were obtained from the Illumina 450k hg19 annotation file (using IlluminaHumanMethylation450kanno.ilmn12.hg19 annotation package in R). Preprocessed methylation intensities were converted to β values and subsequently transformed into M-values, which more closely approximate the assumptions of parametric statistical models^69^. Unless otherwise stated, all analyses were conducted using M values.

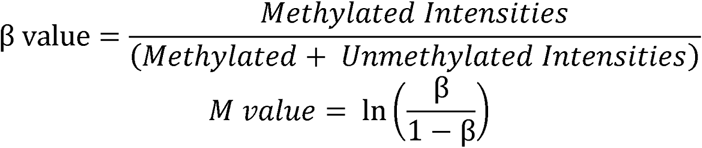

Methylation profiles for the AECM validation cohort were generated using the HELP-tagging assay, which produces paired signal intensities from digestion with a methylation-sensitive enzyme (HpaII) and a methylation-insensitive enzyme (MspI). For each probe, the ratio of HpaII to MspI intensities can be visualized as a point in two-dimensional space, and the angle between this point and the x-axis reflects the degree of methylation^64^. Angles range from 90° (fully unmethylated) to 0° (fully methylated) and were scaled to fall between 0 and 1, serving as pseudo-β values. These values were subsequently logit-transformed into pseudo-M values to facilitate comparability with the TARGET 450k dataset.

### Variably Methylated Region (VMR) Derivation

To reduce noise and redundancy, single-CpG measurements (β values) were aggregated into variably methylated regions (VMRs) using DMRcate (v3.2.1)^42^. Regions were defined based on genomic proximity (λ=500) and then filtered to retain those with high inter-sample variability and within-region concordance. Methylation values within said regions were aggregated into a single value by averaging β values across all CpG sites. CpG island context (island, shore, shelf, open sea), termed resort type, were annotated using the Illumina 450k hg19 annotation file. AECM HELP-tagging methylation probes were then aligned to the TARGET-defined VMRs. Probes that overlapped or fell within 1,000 base pairs of the VMR’s boundaries were averaged into a single aggregate methylation value for each VMR, which we collectively called the AECM VMR set.

### Enhancer Annotations

Due to their tissue-specific nature^70–72^, we sought to identify osteosarcoma-relevant enhancers. To do this, we leveraged H3K27ac ChIP-seq data, a histone marker strongly associated with active enhancers^73^. We analyzed publicly available H3K27ac profiles from localized osteosarcoma bone tumors (GSE234997), generated by a research group at Texas A&M^46^. Utilizing these independent profiles, we applied BEDtools (RRID:SCR_006646, v2.31.1) to identify replicated peaks consistently observed across all three samples, which we designated as osteosarcoma-specific enhancer regions^74^. VMRs overlapping these replicated H3K27ac peaks were classified as enhancer linked. Enhancer-linked VMRs were further stratified by resort type.

### Unsupervised Clustering Analysis

Unsupervised clustering analysis was performed on several profiles using standard workflows. For the global VMR profile in the TARGET cohort, clustering was performed using Uniform Manifold Approximation and Projection (UMAP, umap-learn v0.5.7, n_neighbors = 8 and min_dist = 0.01) for dimensionality reduction in combination with k-means clustering (scikit-learn v1.6.1, n_clusters = 2) applied to the UMAP embeddings^75^.

To assess whether resort-type context influenced clustering, VMRs were stratified into four groups (island, shore, shelf, and open sea) according to Illumina 450k hg19 annotations. VMRs were designated island, shore, shelf, or open sea if 90% or more of its CpG sites belong to the corresponding resort type. Within each subgroup, unsupervised clustering was performed using dimensionality reduction (UMAP) followed by density-based spatial clustering of applications with noise (DBSCAN, v1.2.2)^76^. DBSCAN parameters varied based on subgroup.

VMRs overlapping replicated H3K27ac peaks (see Section 4) were analyzed separately using the same techniques as the resort-type-specific VMR profiles (UMAP + DBSCAN). For consistency, enhancer-linked VMRs were also stratified by resort-type context, and the same clustering approach was applied within each subgroup.

### Supervised Prognostic and Predictive Analyses

Univariable survival analysis was conducted on the TARGET global VMR profile testing for associations with RFS using a Cox proportion hazards (CPH) regression model (lifelines v0.28.0)^77,78^. P-values were adjusted for multiple testing error using Benjamini-Hochberg false discover rate (BH FDR)^79^ correction and were considered significant at q < 0.05.

VMRs were also used to calculate a prognostic signed-average value, as we and others have previously reported^58,59,80–83^. The signed average is calculated for each sample by taking the sign of the direction of the univariable VMR association with RFS and multiplying this across the methylation values for that VMR and then subsequently calculating the average across the given sample. To avoid bias and overfitting, all CPH prognostication models were evaluated using a LOOCV (leave-one-out cross validation) framework^84^, calculating a risk score for the left-out sample at each iteration. Univariable survival analyses and signed-average scores are re-calculated at each iteration. This supervised framework was conducted with and without the utilization of metastasis at diagnosis as a covariate during fitting and application of the model within the LOOCV loop. Risk groups were defined by splitting samples at the median risk score. Model performance was evaluated using the time-dependent receiver operating characteristics (tdROC) method^85^. We further performed 100 permutations of the survival labels and re-ran the cross-validation supervised signed-average modeling pipeline for each permutation. Within each, the number of significant VMRs identified per fold was recorded and summarized across folds. Folds with zero significant VMRs were noted as they preclude further model construction and downstream performance assessment.

Chemoresponse status was available for only a subset of the TARGET cohort and was treated as a binary endpoint (percent necrosis >90%). For patients with available chemoresponse data, an univariable analysis was conducted on the TARGET global VMR profile testing for associations with response to chemotherapy (chemoresponse) by using logistic regression. P-values were adjusted for multiple testing error using BH FDR correction and were considered significant at q < 0.05.

Three supervised models were trained and tested under LOOCV using two distinct feature input strategies. First, a logistic regression model was evaluated using a single signed average value. In parallel, L1-penalized logistic regression (LASSO) and random forest models were evaluated using individual VMRs as distinct feature inputs. Model performance was assessed using ROC^86^ analysis (results quantified by AUC) and supplemented by accuracy-based metrics (available in supplement). Apart from the sparsity intrinsic to L1 regularization, no explicit feature filtering was applied for the primary chemoresponse analyses, however, due to the large number of significant variables, sensitivity analysis was conducted at nominal p-values of 0.005 and 0.0025.

Supervised prognostic models trained on the TARGET cohort were applied to the AECM validation cohort. Importantly, the AECM dataset did not provide patient censoring information, and thus the prognostic model was tested using logistic regression on binarized 5-year survival endpoints. Signed average scores were recalculated for the AECM samples using the direction of the logistic regression coefficients derived from the TARGET dataset. Model performance was once again assessed and quantified by ROC and AUC analyses, respectively.

### Gene Set Enrichment Analysis

To identify functional pathways assocaited with RFS, we performed GSEA using MSigDB collections. Genes mapping to the 61-VMR profile were ranked using a composite score defined as *sign (β) × -log_10_(p)* where *β* represents the Cox regression coefficient and *p* represents the p-value for the association between methylation and RFS. Enrichment was assessed using a weighted Kolmogorov-Smirnov statistic, with significance defined by the Normalized Enrichment Score (NES) and False Discovery Rate (FDR) q-value.

miRNA functional enrichment analysis was performed using miRPath v4.0 (RRID:SCR_017354, DIANA-tools). Predicted miRNA targets were retrieved via the mircoT-CDS database (threshold = 0.7) using miRbase v22.1 annotations. Pathway union significance was characterized by a p-value threshold < 0.05.

### Epigenetic Clock Analysis

Epigenetic age was estimated with a pan-tissue methylation clock originally described by Horvath^48^ and implemented via the *pyaging* package^87^. ΔAge was defined as predicted epigenetic age – chronological age at diagnosis. Association between Δage and chemoresponse was analyzed via t-test. Survival comparison between median-split Δage groups was done via Kaplan-Meier (KM) analysis with significance assessed via log-rank test. Agreement between median-split Δage groupings and global VMR clusters was assessed using χ² test, and relative strength was interpreted using an Adjusted Rand Index (ARI). Finally, risk of recurrence with respect to Δage was assessed using a CPH model^77,78^.

### Comparative analysis of multiple omics layers

RNA-seq counts for the TARGET cohort were normalized using the regularized log (rlog) transformation implemented in DESeq2 (v1.46.0, RRID:SCR_015687)^88^. The rlog transformation stabilizes variance across the dynamic range of expression and was applied prior to downstream clustering. miRNA expression data were generated by qRT-PCR and normalized using the 2^−ΔCt^ method relative to the reference control. Segment-level copy number data was interpreted using GISTIC2.0^89^, and results were further log-transformed and z-score normalized. Normalized gene and miRNA expression as well as CN information were used directly in downstream analyses. For both RNA-seq and miRNA expression profiles, features were ranked and filtered by relative variability using the median absolute deviation (MAD) for RNA-seq and standard variance for miRNA profiling. Unsupervised hierarchical clustering was performed using Euclidean distance and Ward linkage methods. Additionally, all clustering methods were evaluated at 2-, 3-, and 4-cluster partitions. To assess similarity among clustering solutions across all four platforms (methylation, RNA-seq, miRNA profiling, and genomic CN), we calculated the ARI between each pair^90–92^. The ARI measures agreement between two clustering approaches while correcting for random chance. This value ranges from −1 (complete discordance) to 1 (perfect concordance).

### General Statistical Analyses

Survival outcomes were evaluated using (KM) survival analysis^93^. Survival differences between two-cluster solutions were evaluated using log-rank tests, with multi-group log-rank tests being applied when k > 2 groups were compared and stratified log-rank tests in instances where analyses are stratified by an additional covariate^94^. Median survival times, hazard ratios, and p-values were then reported. Hazard ratios were estimated using CPH models^77,78^. Binary associations (e.g., chemoresponse) were tested using χ² or Fisher’s exact tests as appropriate. Two-sided 95% confidence intervals are reported for effect-size estimates, Hazard Ratios, Odds Ratios, and AUCs. Multiple testing error was addressed using BH FDR method, which was applied to both survival and chemoresponse analyses.

### Reproducibility and Code Availability

All analyses were conducted using **R** (v4.4.3, RRID:SCR_001905) and **Python** (v3.11.6, RRID:SCR_008394). In R, commonly used packages included *minfi*^65^, *DMRcate*^42^, *DESeq2*^88^, *timeROC*^95^, and *survival*^96,97^. In Python, core packages included *scikit-learn*^98^, *scipy*^99^, and *lifelines*^77^. A complete list of 43 packages and version numbers is provided in Supplementary Data 17. Illumina 450k methylation array probe annotations were obtained from the IlluminaHumanMethylation450kanno.ilmn12.hg19 package. Where applicable, scripts and workflows were run with fixed seeds to ensure reportability of clustering and modeling. Comprehensive documentation and descriptions for all scripts used throughout this manuscript are provided in a dedicated code reference notebook (Supplementary Data 18) with an active repository available on <u>GitHub</u> to host any future script updates or maintenance.

## Supporting information

Supplementary Data Key

## Data Availability

The datasets analyzed in this study are available the NCI Genomic Data Commons (https://portal.gdc.cancer.gov/projects/TARGET-OS) and the Gene Expression Omnibus repository (GSE59200). All other data are available from the corresponding author upon reasonable request.

https://www.ncbi.nlm.nih.gov/geo/query/acc.cgi?acc=GSE59200

## Acknowledgements

The results published here are in part based upon data generated by the Therapeutically Applicable Research to Generate Effective Treatments (https://ocg.cancer.gov/programs/target) initiative, phs000218. The data used for this analysis are available at https://portal.gdc.cancer.gov/projects. This study was supported by the NIH R01 CA178908 Award to Dimitrios Spentzos, the Amy Chase McMahon Sarcoma Research Fund and the Casper Colson philanthropic donation to Dimitrios Spentzos and the Sarcoma Program at the MGH Cancer Center.

## Corresponding author

Correspondence to Dimitrios Spentzos.

