## Supplementary figures and images for "Epigenetic patterns and methylation-based models for robust outcome prediction in osteosarcoma"

### Supplementary Data 2

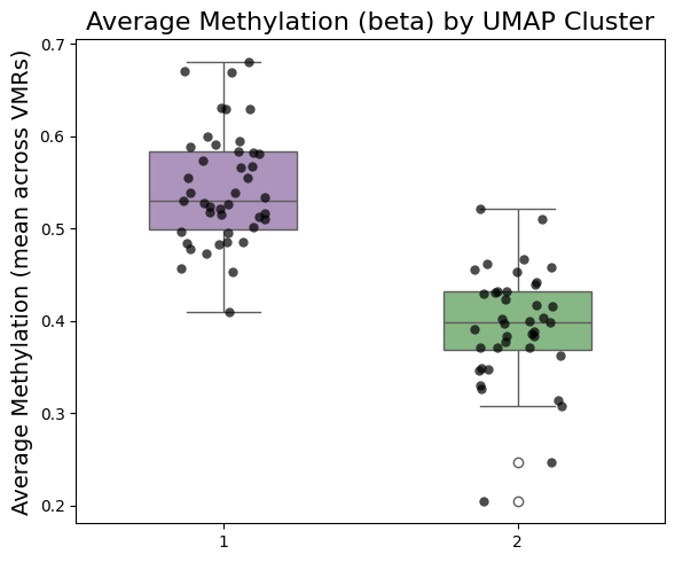

### Supplementary Data 3

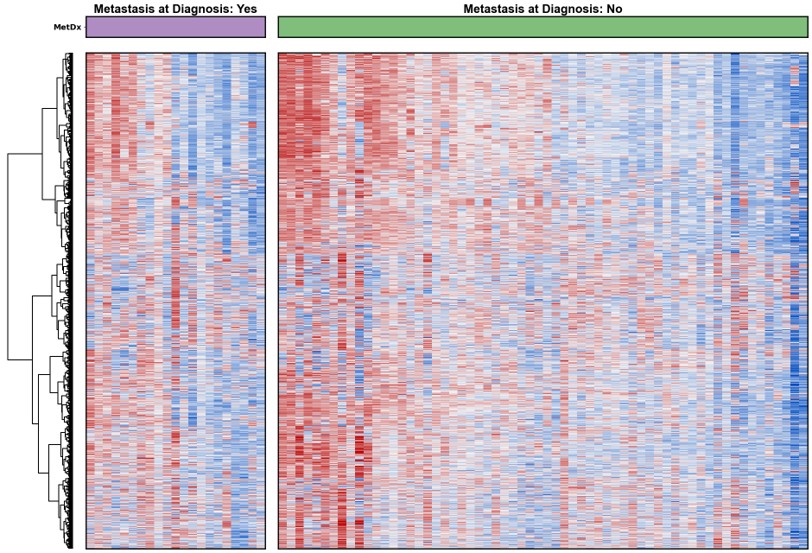

### Supplementary Data 5a

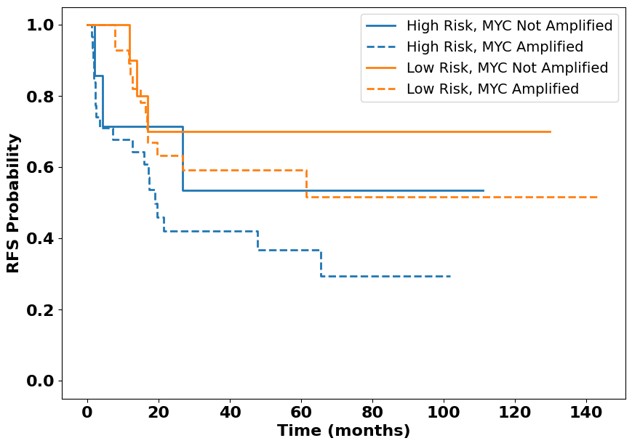

### Supplementary Data 5b

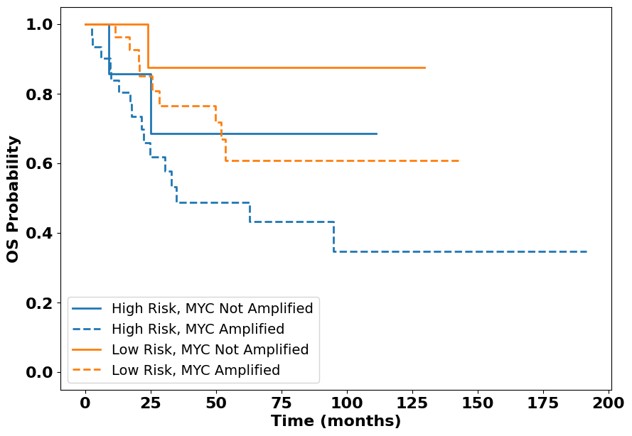

### Supplementary Data 14

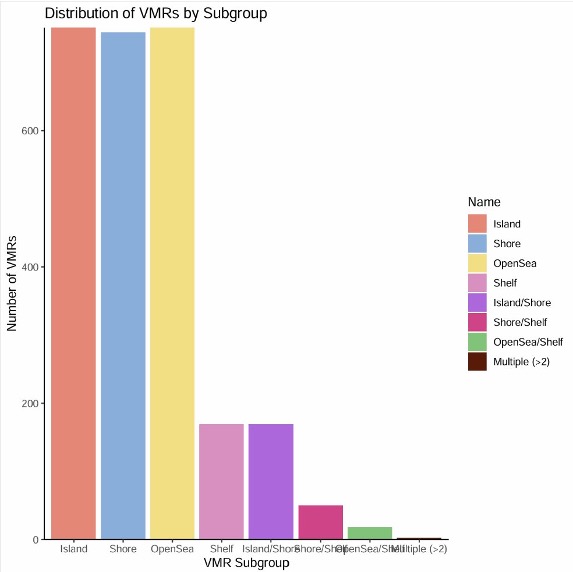

### Supplementary Data 15

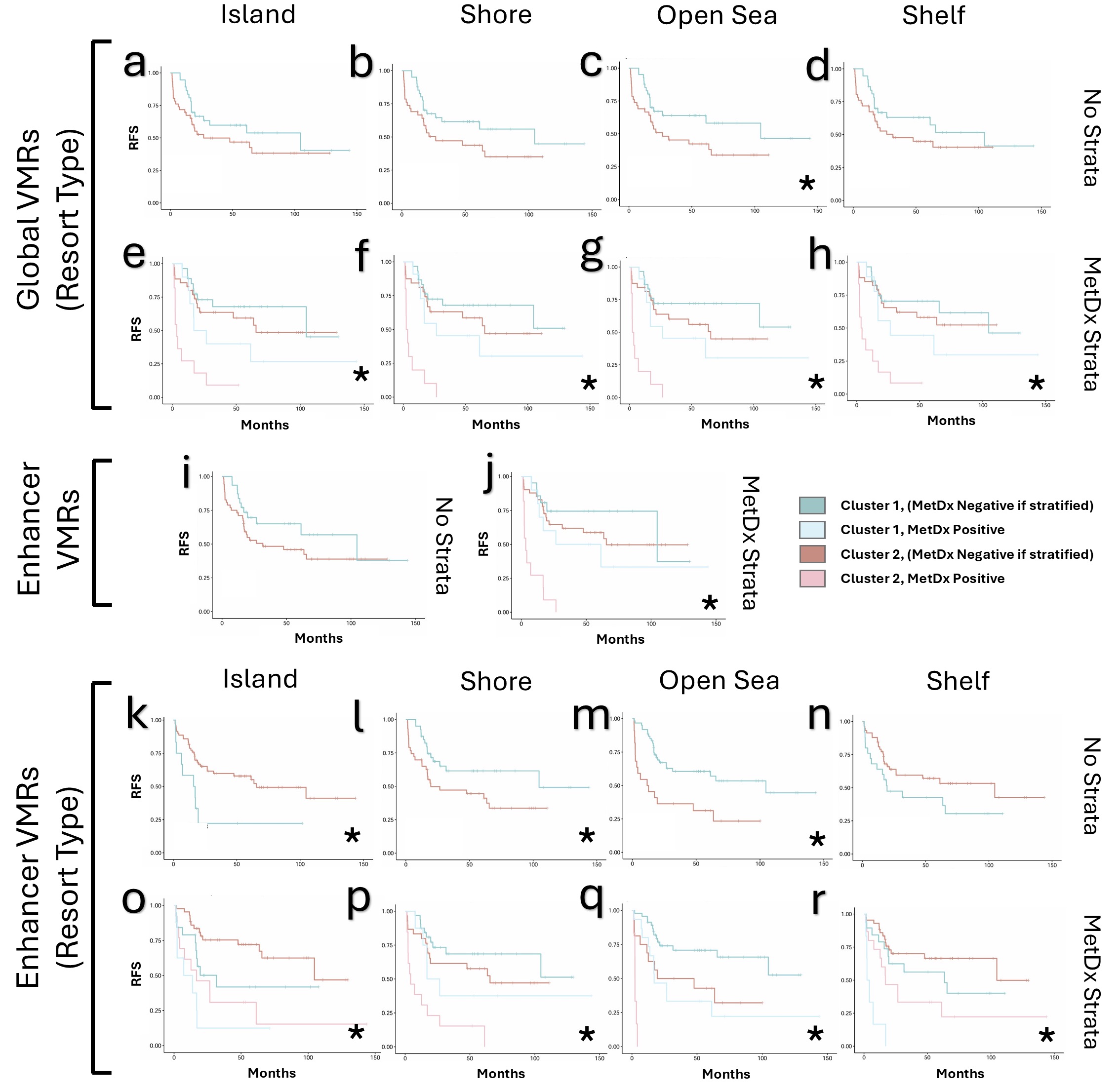

### Supplementary Data 16a

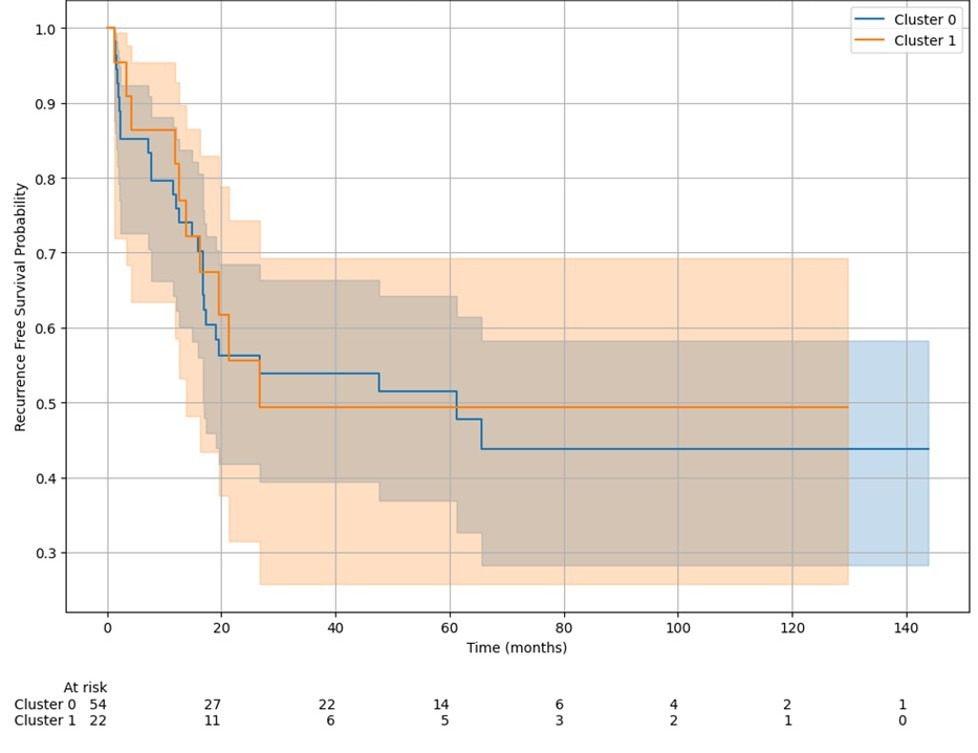

### Supplementary Data 16b

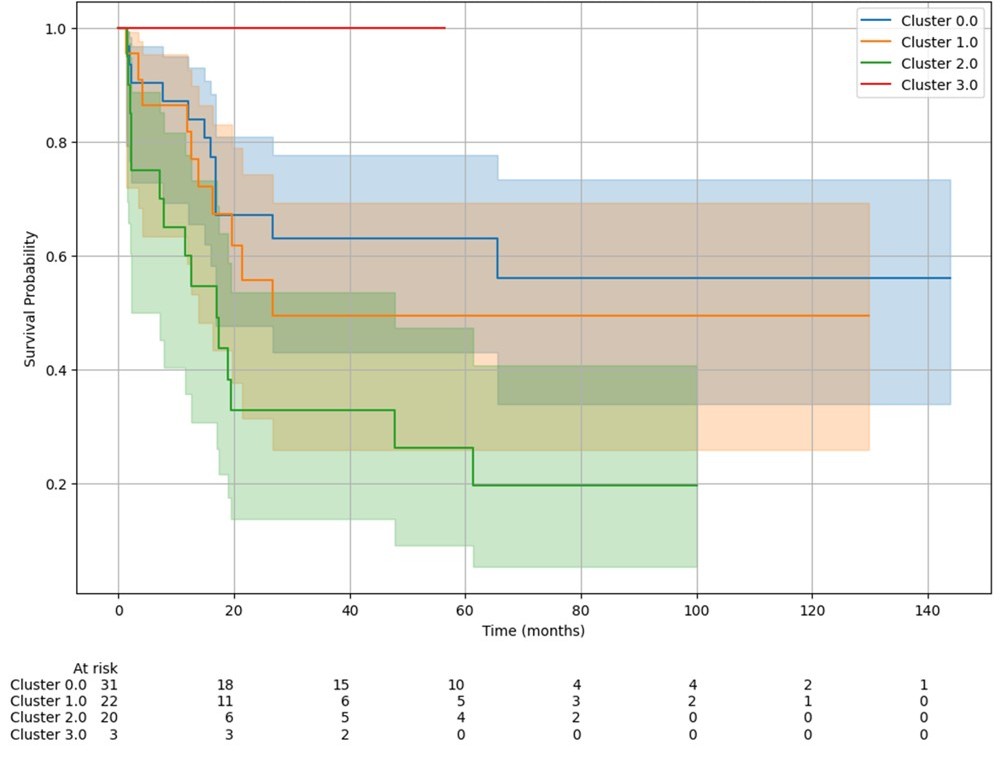
