## Supplementary Data 17 for "Epigenetic patterns and methylation-based models for robust outcome prediction in osteosarcoma"

This is a list of packages/libraries used in R and Python scripts/notebooks over the course of this project.  

## Python v3.11.6

- pandas v2.1.3
- numpy v1.26.2
- matplotlib v3.8.2
- statsmodels v0.14.1
- scikit-learn v1.6.1
- lifelines v0.28.0
- requests v2.31.0
- Bio (from biopython) v1.6.0
- os (stdlib)
- re (stdlib)
- time (stdlib)

## R v4.4.3

- dplyr v1.1.4
- openxlsx v4.2.8
- data.table v1.17.8
- readr v2.1.5
- scales v1.4.0
- Polychrome v1.5.4
- dendextend v1.19.1
- ggplot2 v3.5.2
- RColorBrewer v1.1-3
- GenomeInfoDb v1.42.3
- minfi v1.52.1
- factoextra v1.0.7
- cluster v2.1.8.1
- gplots v3.2.0
- umap v0.2.10.0
- dbscan v1.2.2
- uwot v0.2.3
- dendsort v0.3.4
- pheatmap v1.0.13
- amap v0.8-20
- tidyverse v2.0.0
- stats v4.4.3
- timeROC v0.4
- survival v3.8-3
- bumphunter v1.48.0
- DMRcate v3.2.1
- dbplyr v2.5.0
- GenomicRanges 1.58.0
- MVA 1.0-9
- BiocManager (for installation only, not runtime) v1.30.26
    - IlluminaHumanMethylation450kmanifest v0.4.0
    - IlluminaHumanMethylation450kanno.ilmn12.hg19 v0.6.1

---

Note:
- Base R/Python packages (e.g., `os`, `re`, `time` for Python; `stats` for R) are part of standard library and do not require separate installation.
