## Supplementary Data Key for "Epigenetic patterns and methylation-based models for robust outcome prediction in osteosarcoma"

**Supplementary Data 1 – TARGET dataset derived global VMR profile:**

Data file containing 4427 TARGET-derived VMRs defined using DMRcate and their respective beta- and M-values.

**Supplementary Data 2 – Average beta value by UMAP cluster:**

Figure showing a box plot of the average methylation beta value by UMAP cluster as represented as the mean across all VMRs for a given sample.

**Supplementary Data 3 – Global VMR heatmap according to the presence of Metastasis at Diagnosis (MetDx):**

Figure showing a heatmap of the methylation (M-value) across all VMRs in the global VMR profile. Samples grouped according to the presence or not of Metastasis at Diagnosis.

**Supplementary Data 4 – Unsupervised cluster and supervised risk group association analysis for clinicopathologic and molecular variables.**

Data file containing an unsupervised cluster and supervised risk group association analysis that tests an array of potential confounders.

**Supplementary Data 5 – MYC-stratified risk group survival analysis:**

Figures showing MYC-stratified unsupervised risk group Kaplan-Meier survival analyses for recurrence-free survival (5a) and overall survival (5b) end points.

**Supplementary Data 6 – Univariable VMR survival analysis:**

Data file containing univariable Cox Proportional Hazards VMR survival analysis for both recurrence-free survival and overall survival end points.

**Supplementary Data 7 – Univariable VRM survival analysis corrected for Metastasis at Diagnosis (MetDx):**

Data file containing MetDx-corrected univariable Cox Proportional Hazards VMR survival analysis for both recurrence-free survival and overall survival end points.

**Supplementary Data 8 – Methylation-based GSEA analysis:**

Data file containing a methylation-based GSEA analyses across multiple MSigDB collections (Hallmark, C1, C2, C3, C4, C6, C7, C8)

**Supplementary Data 9 – miRNAPath enrichment analysis:**

Data file containing miRPath enrichment analysis for miRNAs identified in methylation-based GSEA. These results include a full list of miRNA targeted biological pathways and their respective significance values across multiple pathway collections.

**Supplementary Data 10 – Supervised TARGET risk scores:**

Data file containing risk scores for all 83 TARGET samples produced by the supervised signed average Cox Proportional Hazards model with an VMR variance filtering FDR q-value stringency of 0.1.

**Supplementary Data 11 – Univariable VMR chemoresponse analysis:**

Data file containing univariable VMR chemoresponse analysis.

**Supplementary Data 12 – Supervised TARGET chemoresponse predictions:**

Data file containing supervised chemoresponse prediction analysis results.

**Supplementary Data 13 – AECM-mapped VMR profile:**

Data file containing a list of 3388 VMRs mapping to at least one AECM methylation site.

**Supplementary Data 14 – Distribution of VMRs by resort type:**

Figure showing the distribution of the global TARGET VMR profile by their respective resort type functional subgroups.

**Supplementary Data 15 – Functional group analysis in the TARGET dataset:**

Figure showing the breakdown of survival differences amongst the various functional group subsets in the TARGET dataset. This includes a stratification for enhancer-linked VMRs.

**Supplementary Data 16 – TARGET copy number survival analysis:**

Figures showing the Kaplan-Meier survival analysis of 2- (16a) and 4-cluster (16b) solutions in TARGET copy number dataset.

**Supplementary Data 17 – Computational Version History:**

A list of base packages/libraries used in R and Python scripts throughout the manuscript.

**Supplementary Data 18 – Code Reference Notebook:**

A Jupyter notebook containing a comprehensive breakdown of all code utilized throughout the manuscript.
