## Supplementary Data PDF for "Epigenetic patterns and methylation-based models for robust outcome prediction in osteosarcoma"

**Supplementary Data 2:**

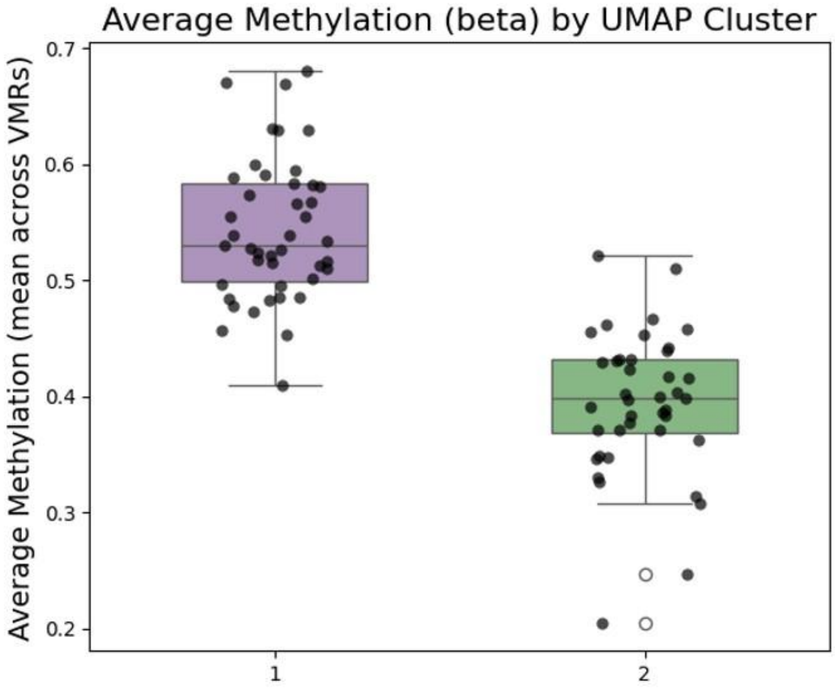

**Supplementary Data 3:**

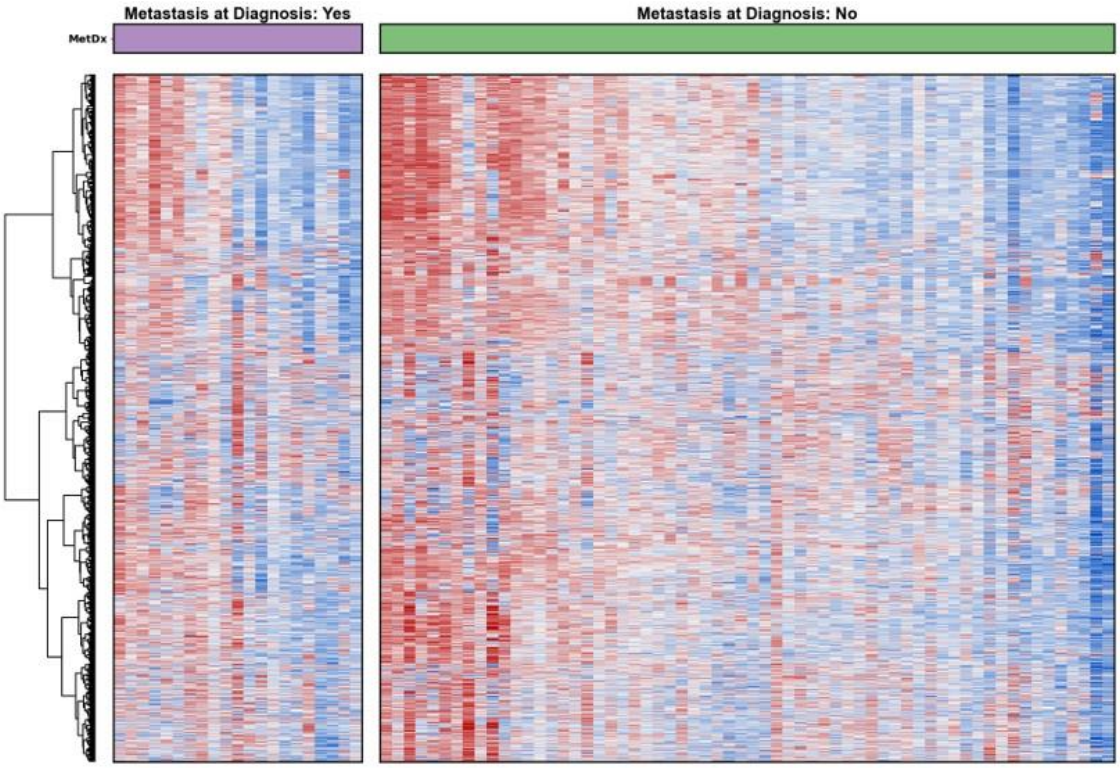

**Supplementary Data 4:**

| Variable | Test | Statistic | p-value |
|---|---|---|---|
| Metastasis | Chi2 | 0.0368 | 0.8479 |
| Age (Cont) | t-test | 0.3258 | 0.7454 |
| Age (Bin) | Chi2 | 0.013 | 0.9094 |
| Sex | Chi2 | 0.0045 | 0.9468 |
| Batch | Chi2 | 2.635 | 0.4514 |
| TERT Expr | Chi2 | 0.0538 | 0.8166 |
| ATRX Expr | Chi2 | 0.7151 | 0.3978 |
| ATRX Mutation | Chi2 | 1.681 | 0.1948 |
| TP53 Mutation | Chi2 | 0.4415 | 0.5064 |
| MDM2 Mutation | Chi2 | 0 | 1 |
| RB1 Mutation | Chi2 | 1.3525 | 0.2448 |
| CDK2A Mutation | Chi2 | 0 | 1 |
| chemoresponse | Chi2 | 15.4523 | 0.0004 |
| myc | Chi2 | 0.3031 | 0.582 |

**Supplementary Data 5a/b:**

a

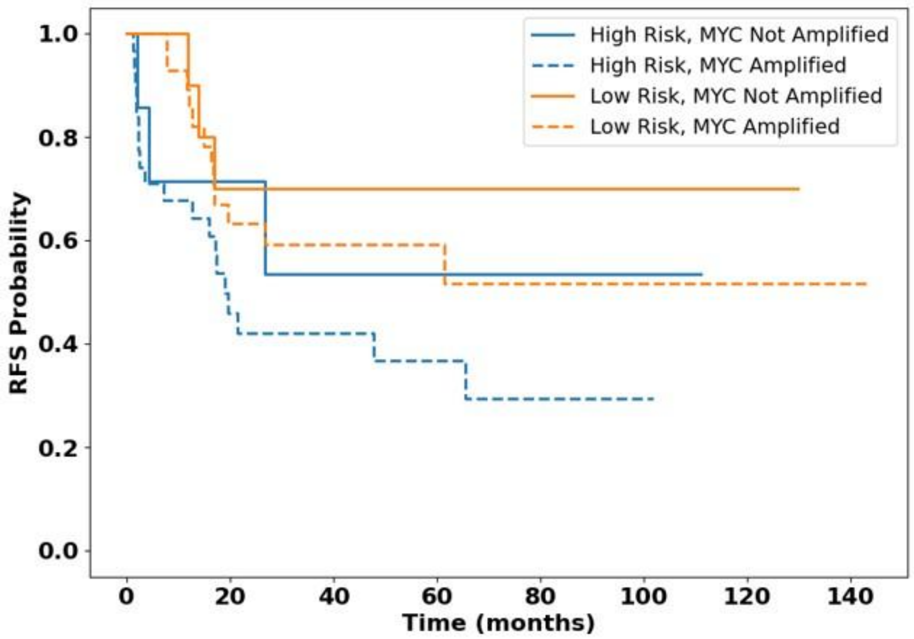


b

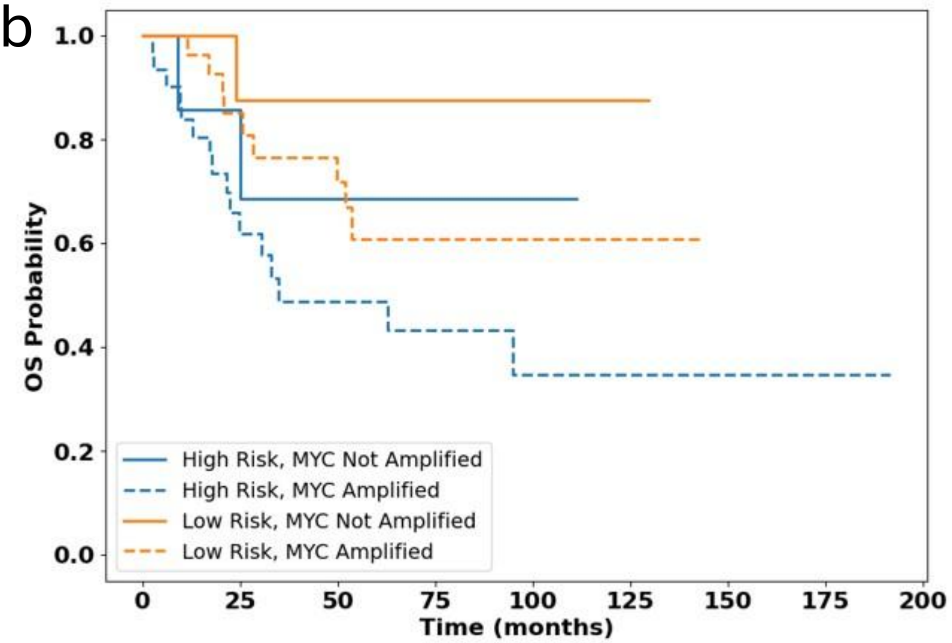

**Supplementary Data 14:**

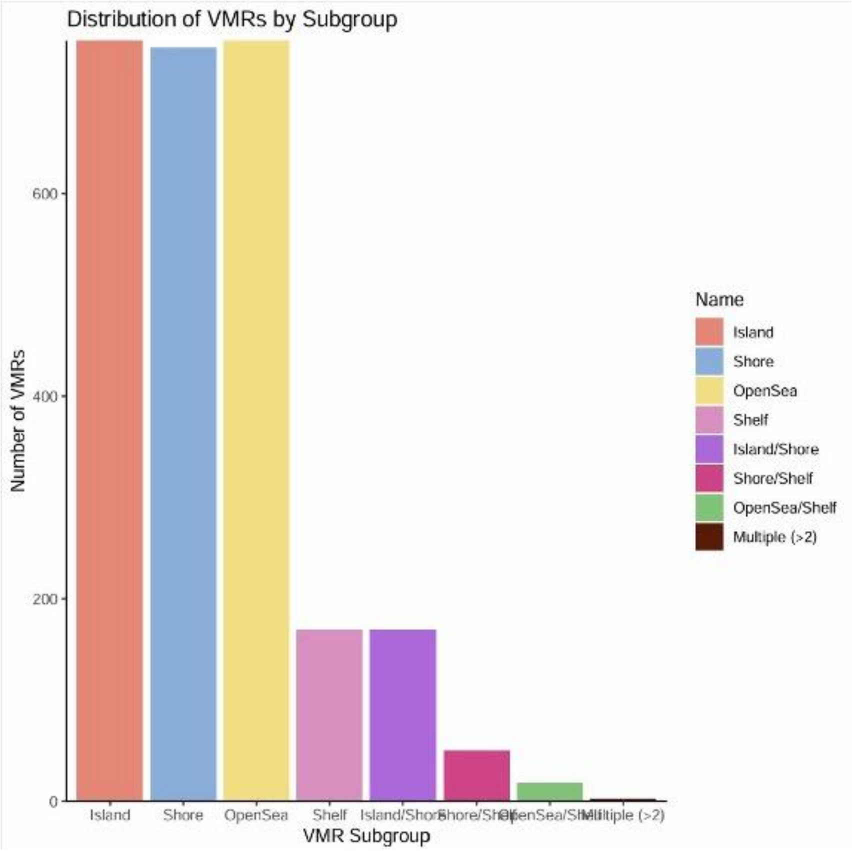

**Supplementary Data 15:**

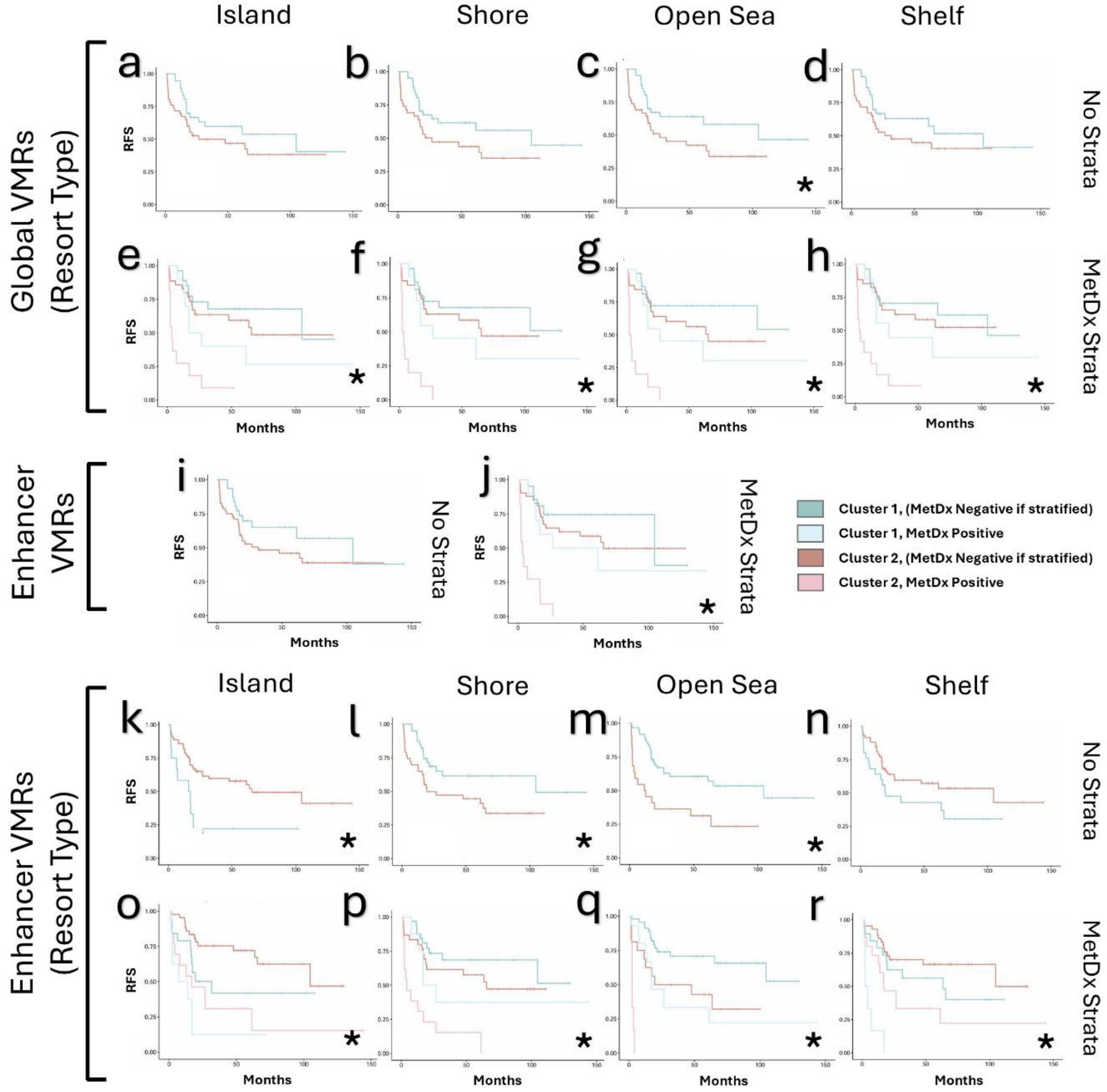


**Supplementary Data 15: Functional group analysis in the TARGET dataset**

a-d: RFS analysis of global profile, across resort subgroups (log-rank p = 0.160, 0.065, 0.042, 0.160). Figures 2e-h: MetDx stratified RFS analysis of global profile, across resort subgroups (multi-group log-rank p = 0.040, 0.007, 0.004, 0.050). Supplementary Data 13i: RFS analysis of enhancer-associated VMRs (log-rank p = 0.14). j: MetDx stratified RFS analysis of enhancer-associated VMRs (multi-group log-rank p = 0.009). k-n: RFS analysis of enhancer-associated VMRs, across resort subgroups (log-rank p = 0.001, 0.029, 0.001, 0.087). o-r: MetDx stratified RFS analysis of enhancer-associated VMRs, across resort subgroups (multi-group log-rank p = 0.050, $2.1 \times 10^{-5}$, 0.017, 0.031)

*Note: * denote p <= 0.05*

**Supplementary Data 16:**

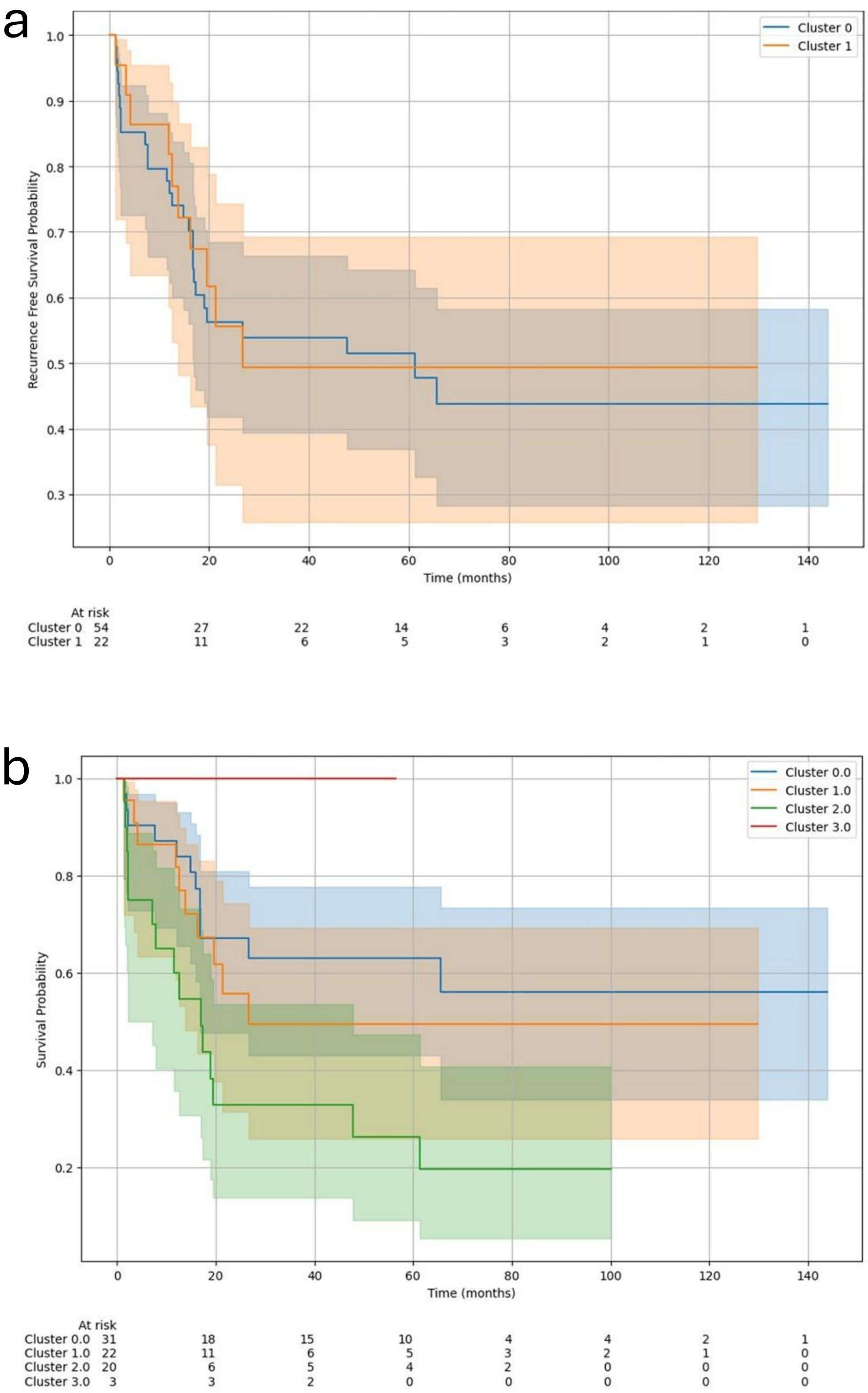
